# Deep Learning and Holt-Trend Algorithms for predicting COVID-19 pandemic

**DOI:** 10.1101/2020.06.03.20121590

**Authors:** Theyazn H.H Aldhyani, Melfi Alrasheed, Ahmed Abdullah Alqarni, Mohammed Y. Alzahrani, Ahmed H. Alahmadi

**Affiliations:** Department of Computer Sciences, King Faisal University, Saudi Arabia; Department of Quantitative Methods, School of Business, King Faisal University. Saudi Arabia; Department of Computer Sciences and Information Technology, Albaha University, Saudi Arabia; Department of Computer Science and Information at Taibah University, Kingdom of Saudi Arabia

**Keywords:** Deep learning algorithm, Holt-Trend model, Prediction COVID-19, machine learning

## Abstract

According to WHO, more than one million individuals are infected with COVID-19, and around 20000 people have died because of this infectious disease around the world. In addition, COVID-19 epidemic poses serious public health threat to the world where people with little or no pre-existing human immunity can be more vulnerable to the effects of the effects of the coronavirus. Thus, developing surveillance systems for predicting COVID-19 pandemic in an early stage saves millions of lives. In this study, the deep learning algorithm and Holt-trend model is proposed to predict coronavirus. The Long-Short Term Memory (LSTM) algorithm and Holt-trend were applied to predict confirmed numbers and death cases. The real time data have been collected from the World Health Organization (WHO). In the proposed research, we have considered three countries to test the proposed model namely Saudi Arabia, Spain and Italy. The results suggest that the LSTM models showed better performance in predicting the cases of coronavirus patients. Standard measure performance MSE, RMSE, Mean error and correlation are employed to estimate the results of the proposed models. The empirical results of the LSTM by using correlation metric are 99.94%, 99.94% and 99.91 to predict number of confirmed cases on COVID-19 in three countries. Regarding the prediction results of LSTM model to predict the number of death on COVID-19 are 99.86%, 98.876% and 99.16 with respect to the Saudi Arabia, Italy and Spain respectively. Similarly the experimented results of Holt-Trend to predict the number of confirmed cases on COVID-19 by using correlation metrics are 99.06%, 99.96% and 99.94, whereas the results of Holt-Trend to predict the number of death cases are 99.80%, 99.96 and 99.94 with respect to the Saudi Arabia, Italy and Spain respectively. The empirical results indicate the efficient performance of the presented model in predicting the number of confirmed and death cases of COVID-19 in these countries. Such findings provide better insights about the future of COVID-19 in general. The results were obtained by applying the time series models which needs to be considered for the sake of saving the lives of many people.

## 1. Introduction

Currently, The COVID-19 pandemic is regarded as a global outbreak threat for the global health. Coronaviruses are large species of viruses that may cause diseases to animals and humans. A number of coronaviruses are known to cause respiratory infections in humans, ranging from common colds to more severe diseases such as Middle East Respiratory Syndrome (MERS) and Severe Acute Respiratory Syndrome (SARS). The recently discovered Coronavirus (CORONA) is known as Covid-19 which is an infectious disease caused by the recently discovered Coronavirus [1]. There was no knowledge of the presence of the emerging disease prior to the outbreak of the virus in the Chinese city of Wuhan in December 2019. Figure 1 shows how the SARS-COV, SARS-COV2 and COVID-19 transfer from various animals like Camel, Pig, Cow, Rate and Bat to human. The most common symptoms of COVID-19 are fever, fatigue and dry cough. Some patients may experience pain, nasal congestion, coldness, sore throat, or diarrhea. These symptoms are usually mild and begin gradually. Some people become infected without showing any symptoms and without feeling sick. Most people (about 80%) are recovering of the disease without the need for special treatment [1]. Approximately one in six people with COVID-19 infection are mostly with severe symptoms. People can be infected with Coved-19 through other people living with the virus. The disease can be transmitted from person to person through small droplets that are scattered from the nose or mouth when a person with COVID-19 coughs or sneezes. These droplets fall on objects and surfaces surrounding the person. Other people can then develop COVID-19 disease when they touch these objects or surfaces and then touch their eyes, nose or mouth. People can also develop COVID-19 if they breathe the droplets that come out of the affected person with their cough or exhalation. It is therefore important to stay more than one meter (3 feet) away from the sick person. This health crisis will lead to significant economic repercussions, reflecting shocks to supply and demand that differ from previous crises. Substantial policies need to be developed to help economies overcome the epidemic, while maintaining the integrity of the network of economic and financial relations between workers and businesses, lenders and borrowers, suppliers and end-users, so that activity can recover once the outbreak ends. The aim is to prevent such a temporary crisis from causing permanent harm to people and companies through job losses and bankruptcies. The loss of life from the outbreak of COVID-19 has increased at an alarming rate, while the disease is spreading through larger number of countries. It is clear that the highest priority should be given to maintaining the health and safety of people as much as possible. Countries can help by spending more to support their health systems, including spending on personal protective equipment, testing, diagnostic tests and adding more beds in hospitals. While a vaccine has not yet been found to stop the spread of the virus, countries have taken actions to curb its spread. The economic impact is already evident in the countries which mostly affected by the outbreak. For example, in China, manufacturing and services activities fell sharply in February. While activity in the manufacturing sector has fallen comparable to its level at the beginning of the global financial crisis, the decline in services appears to be even greater this time – due to the significant impact of social distancing [2].

**Figure 1.**
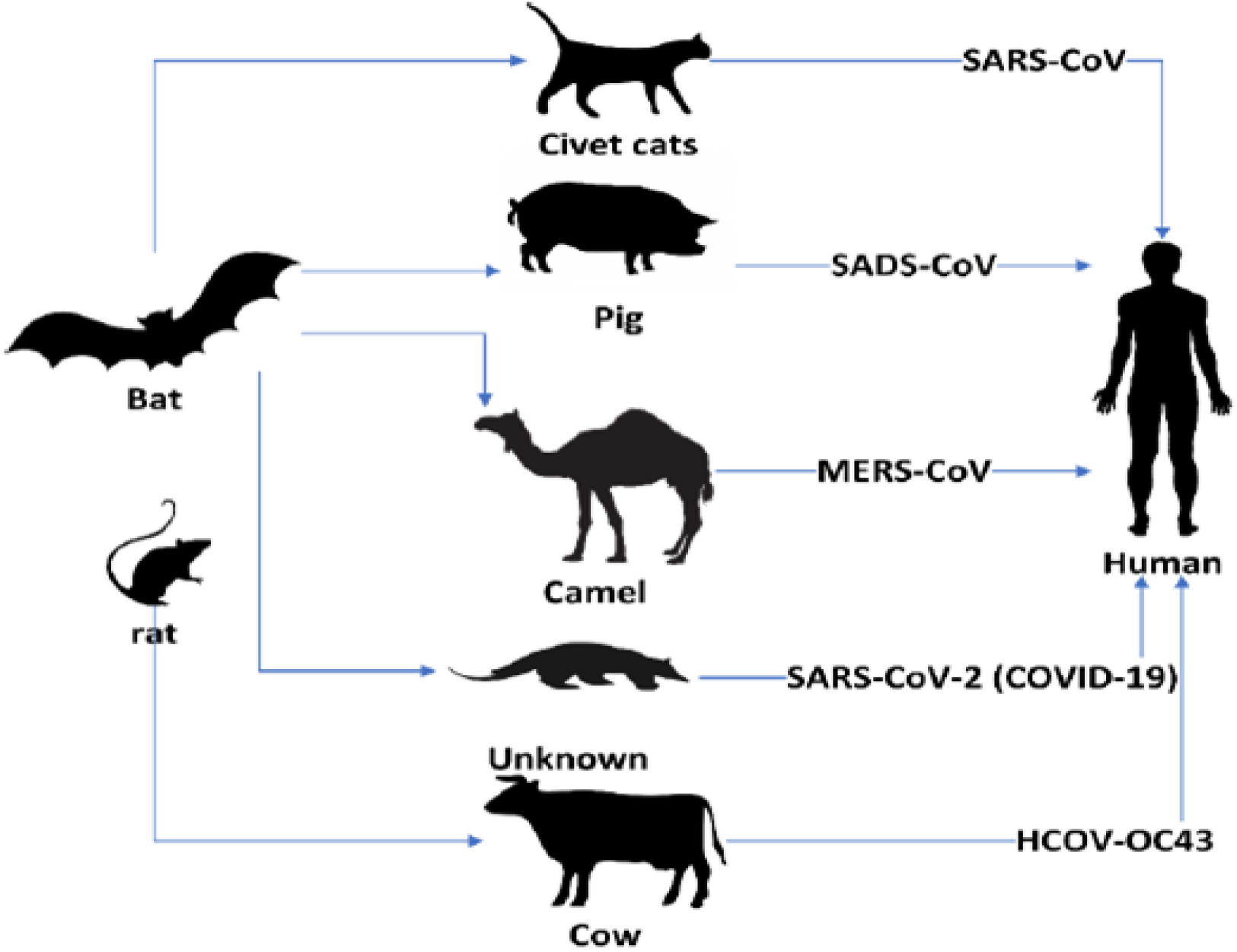
How the COVID 19 transmission from animals to human.

In this study, we have developed models that have the capability to predict the COVID-19 epidemic in the best accuracy. The main contribution of this paper is presenting the LSTM and Holt-Trend models as effective approaches for predicting the numbers of confirmed and death cases in Saudi Arabia, Italy and Spain. The results of time series models to predict COVID-19 epidemic based on real time data that are gathered from WHO were more satisfactory.

Researchers have used the search queries as a potential new health information for surveillance and monitoring heath care system [3,4,5,6–10]. The using social media for prediction disease [11]. The Goggle is one of most search engine, the search terms that searched by numbers of people for discovering specific disease [12,13]. Form January 2004, adjusted total search volume of data from geographic region due to this information carry significant pattern can help to identify any issues [14]. The numbers of researchers used Google trend to identify epidemics of infectious diseases, like influenza 12,13], chickenpox [15], gastroenteritis[16]. [13]. McCarthy [3] used Google Trend data for prediction suicide risk on a population-wide level. Figure 2 shows how to track COVID19 spread over the global.

**Figure 2.**
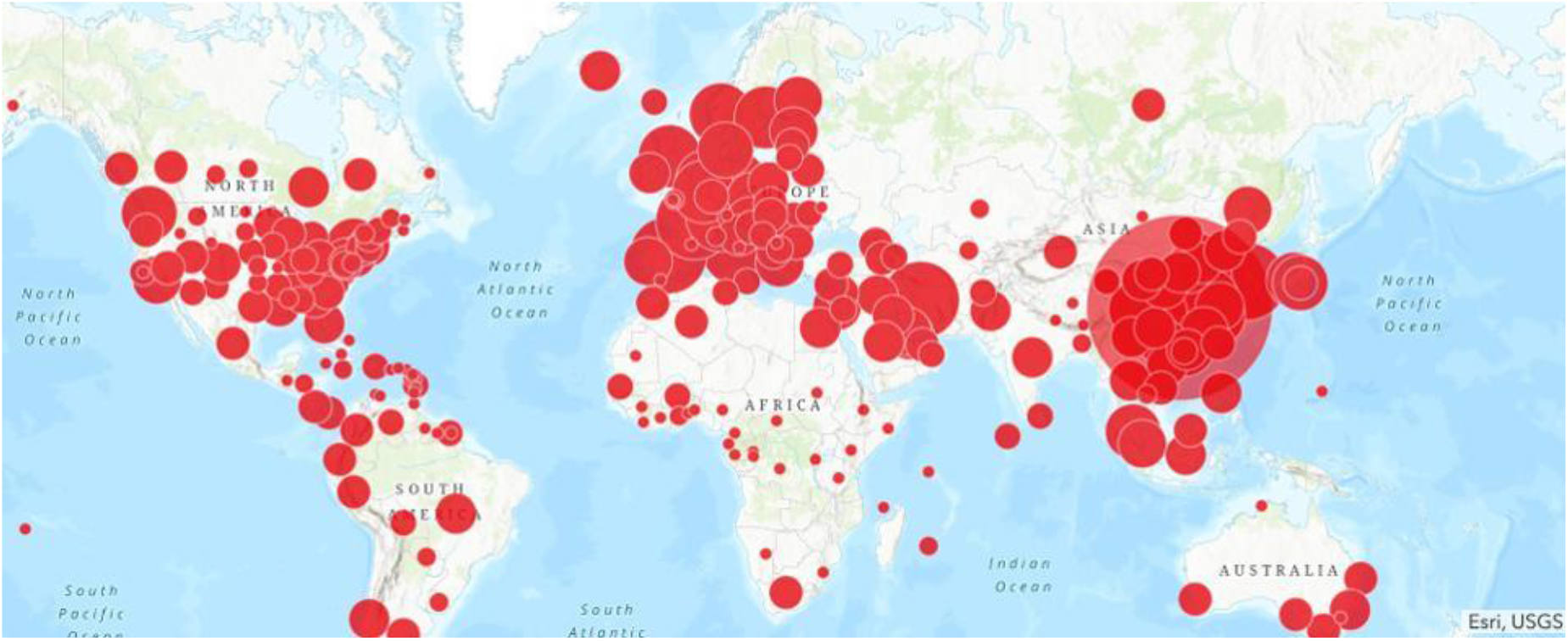
Shows to track COVID19 spread over global.

There are numbers of researchers have been developed models for prediction the infectious disease. Using the Internet search queries to predict infectious disease [17–21]. The internet search data is very important for analyzing and prediction the new epidemic, due to most of the people round the world use the internet for searching the specific information for things happen at the current time, according to Towers et al. Internet search data can help to develop surveillance system for the detection of the epidemic [23]. For instance, Huang et al. proposed generalized additive model (GAM), model to predict hand, foot, and mouth diseases using search internet queries, the results of the proposed approved that the system can detect the disease before spreading. It is observed that the big data surveillance tools have the benefit of being easy to access and can detect infectious disease trends before health officials [24]. Furthermore, the social media is considered as internet source information for predicting and analyzing the outbreak disease. Tenkanen et al. report that social media big data is relatively easy to collect and can be used freely, which means accessibility is satisfactory and the data is created continuously in real time with rich content [25]. Using twitter data to predict mental illness [26] and infectious epidemics [27–29] and can be used for predictions in a variety of other scientific fields [30–33]. Shin et al. presented to predict the infectious diseases with using twitter data, it is noted that there is strong relationship between twitter data and infectious diseases and there are possibility for developing surveillance systems to predict infectious disease using twitter data [27]. These studies used deep learning algorithm to predict infectious disease [34,35]. Deep learning algorithm is used to analyze big data [36]. Deep learning algorithm is more powerful algorithm for prediction pattern from big data, the results of the deep learning are really satisfactory according to [37–39]. According to by Xu et al., the performance of deep learning algorithm is better than generalized linear model (GLM), the least absolute shrinkage and selection operator model, and the autoregressive integrated moving average (ARIMA) model [34]. Aldhyani et al. [40] proposed the ANFIS model to predict chronic dieses using Google trend data. Aldhyani et al. [41]. Presented soft clustering to applied machine learning algorithms to classify the chronic dieses. Min Kang et al., [42], Qingyu Yuan et al., [43], and hilip M et al., [44] developed the surveillance system to predict influenza activity by using internet search queries. Gabriel J. Milinovich et al., [45] presented a framework to estimate infectious diseases in Australia by using internet search queries. They observed that web search activities have a potential role in predicting emerging infectious disease events. Samantha Cook et al., [46] compared their methodology. The objective of the present research was thus to build a model that assists in predicting the COVID-19 epidemic. We have applied advance time series models to predict the cases of COVID-19 epidemic.

## 2. Materials and Methods

This section presents the proposed method for predicting CAVID-19. Figure 3 displays the overall formwork of the proposed model to predict CAVID-19. This research includes real time data that have been collected from WHO for three countries to test the proposed model. The min-max normalization algorithm is used to normalize the data. The deep learning and Holt-Trend models are proposed to predict the number of confirmed and death cases.

**Figure 3.**
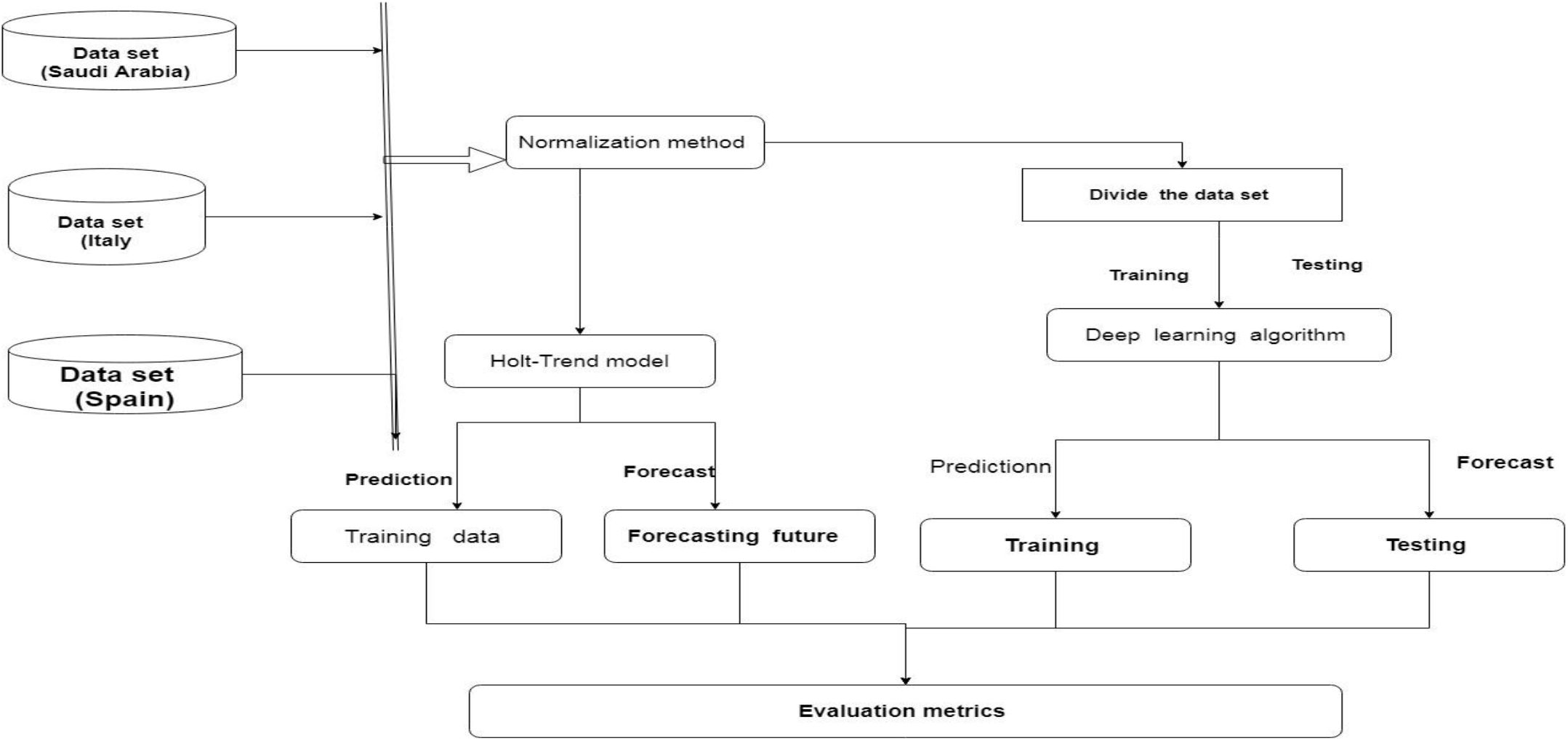
Formwork of proposed model.

### 2.1 Research data

As mentioned above, the data sets have been collected from WHO for three countries namely Saudi Arabia, Italy and Spain. The Data of 85 days, i.e. between 21 January 2020 and 15 April 2020 was used in the current study. In this study, the confirmed cases are considered for predicting future spread of the coronavirus. Table 1 summarizes the real dataset. [1], [47].

**Table 1.**
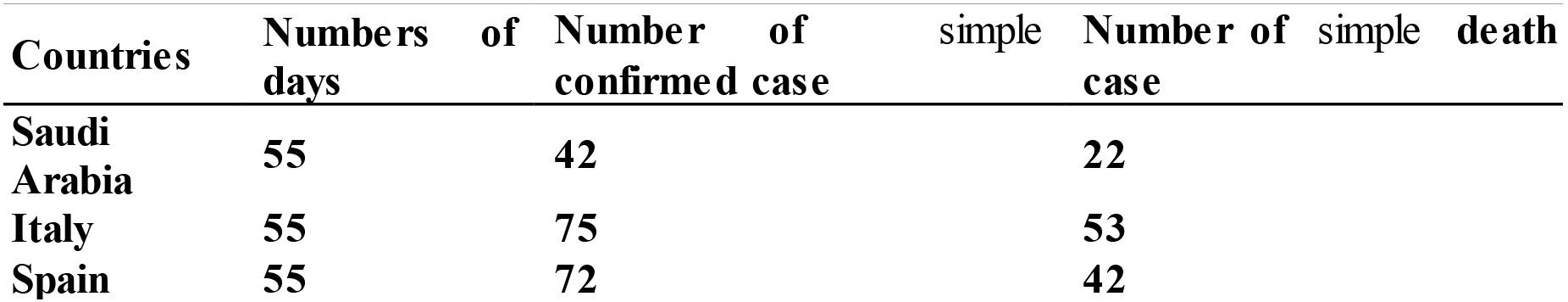
Summarize the real dataset.

### 2.2 **Normalization Method**

Min-max method is employed in Matlab for scaling the data. This method has transformed data within a range of 0 to 1 scales.

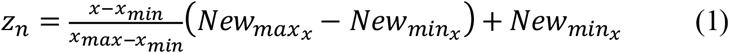

### 2.3 Prediction Models

This section presents the prediction models used to predict the numbers of confirmed and death cases in three countries namely Saudi Arabia, Italy and Spain.

#### 2.3.1 Recurrent Neural Network (RNN)

The Recurrent Neural Network (RNN) was designed in the 1980s [48].The RNN algorithm consists of hidden layers, an input layer and an output layer. The RNN algorithm has a chain like-structure for repeating cells of the RNN algorithm, used to store significant information from previous process steps. Figure **4** shows the structure of LSTM algorithm.

**Figure 4.**
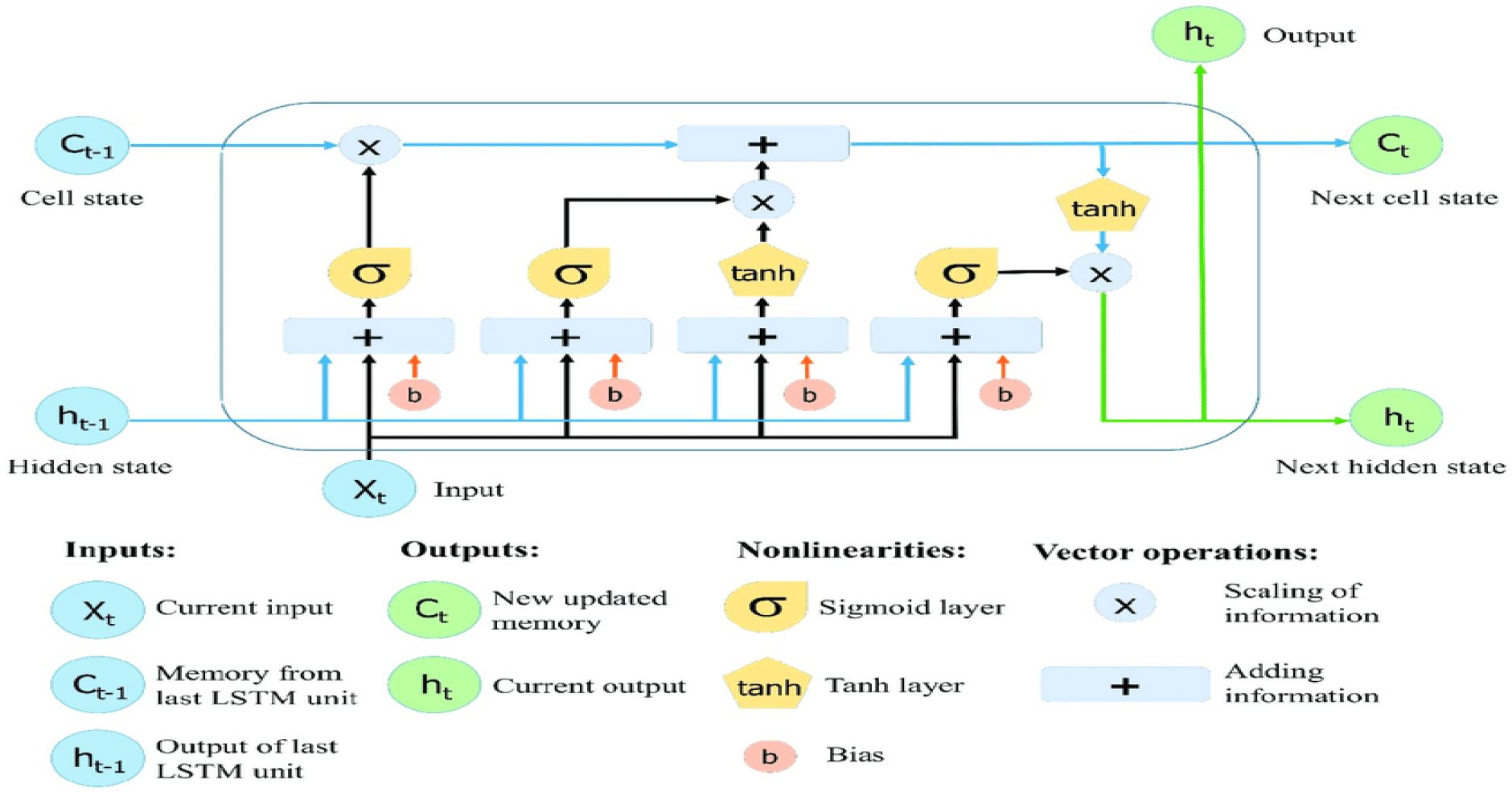
Structure of the LSTM.

The hidden layer is represented by *h_t_* from input *x_t_* to output *y_t_*. Furthermore, the RNN has recurrent loop, which loop back to the past to express that the output is not only a function of new input but also a function of past hidden layer and that way the network keeps growing. The RNN is capable of addressing the issue of exploding and vanishing gradient with using loop, which allow the information to persist. The RNN supports the process of cell state, that helps to transmit the information between the cells with *y_t_* data and embedded on the top of the all.

Figure 5 displays the chunck of neural network. The *x_t_* is the input, whereas the *y_t_* value is the output. If we loop to the figure, the looK value help the information to passed from one step of the network cell to next network cell. The loops create the RNN type of ambiguous object, as we know, the RNN is totally different from normal neural network. The RNN is multiple copies of the same network each passing message to a successor. The cell state in network is a mechanism like conveyor belt, it carries the information through the entire chain. The cells have gates, these gates contains sigmoid function, the output gate value and *y_t_* are subject of multiplication. The sigmoid function can take values between 0 and 1, the 0 value refers to transition information, whereas the value refers 1 close the entire information.

**Figure 5.**
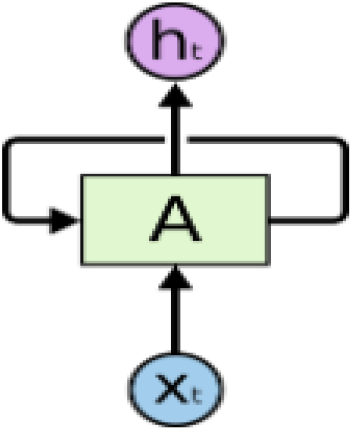
Show chunck of neural network.

Figure 6 shows the unrolled loop, the RNN are shown like sequence and list. Let’s, consider the hidden layer *y_t_* at time step t. the LSTM cell need to decide the cell status

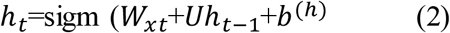

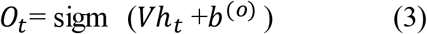

**Figure 6.**
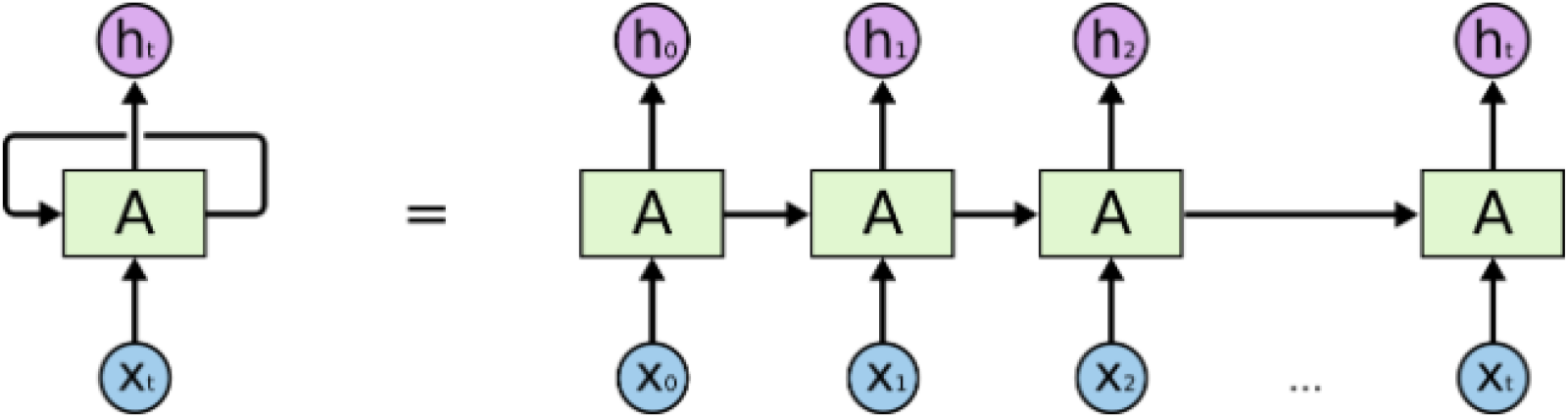
Displays of the unrolled loop of RNN.

Where, *h_t_* is hidden layer correspond to *xt*, *h*_*t*−1_ is hidden state of recurrent neural network, *xt* is input data, *O*_t_ is output value. Where as W, U, and V are the weight vector of neural network, the bias vector of neural network is presented by *b*. In order to transfer the value from hidden layer into the output, the activation function is used. The structure of The long-short term memory cell is shown in Figure 2. It contains forget gate (*f*_*t*_), input gate (it), input modulation gate (*m_t_*), output gate (*O_t_*), memory cell (*c_t_*), and hidden state (*h_t_*). The gates are computed:

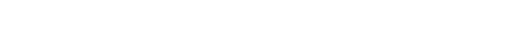

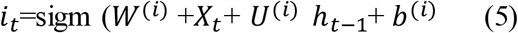

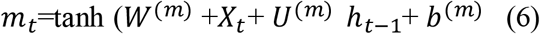

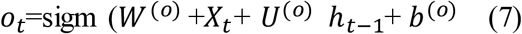

where *x_t_* is an training input data, W and U are parameters used to adjust the weight matrices and *h*_*t*−1_ is the previous hidden layer in The long-short term memory network. In order to transfer the data from input into output by the logistic sigmoid function. The hyperbolic tangent function base on the tanh function, the b is the bias vector of training data. We computed memory cell (*c_t_*) and hidden state (*h_t_*) by these equation:

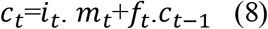

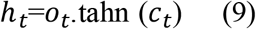

Figure 7 Flow steps of the LSTM model to predict COVID-19. Table 2 demonstrate the significant parameters of the LSTM algorithm to predict COVID-19. It noted the parameters were significant for obtaining the better prediction.

**Table 2.**
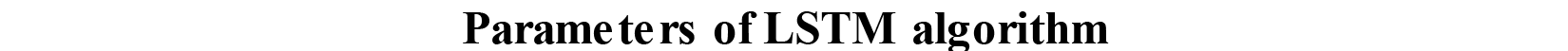
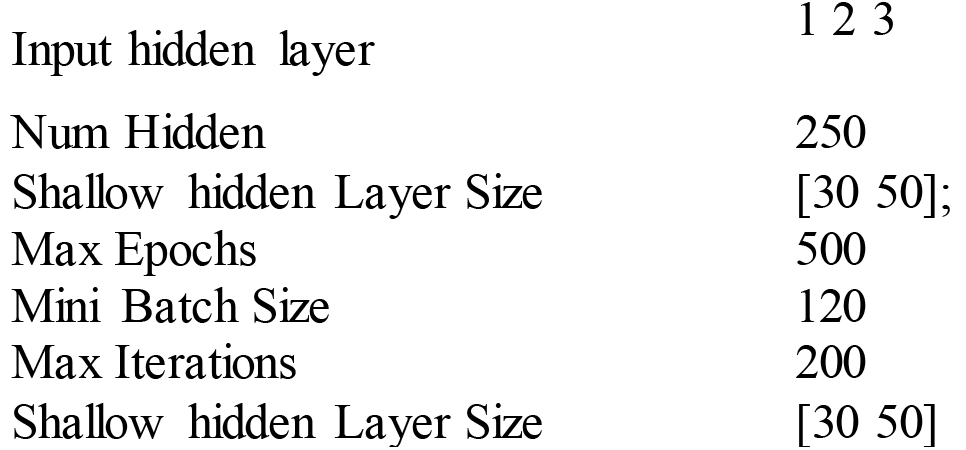
Shows parameters of LSTM algorithm.

|  |  |
| --- | --- |
| Input hidden layer | 1 2 3 |
| Num Hidden | 250 |
| Shallow hidden Layer Size | [30 50]; |
| Max Epochs | 500 |
| Mini Batch Size | 120 |
| Max Iterations | 200 |
| Shallow hidden Layer Size | [30 50] |

**Figure 7.**
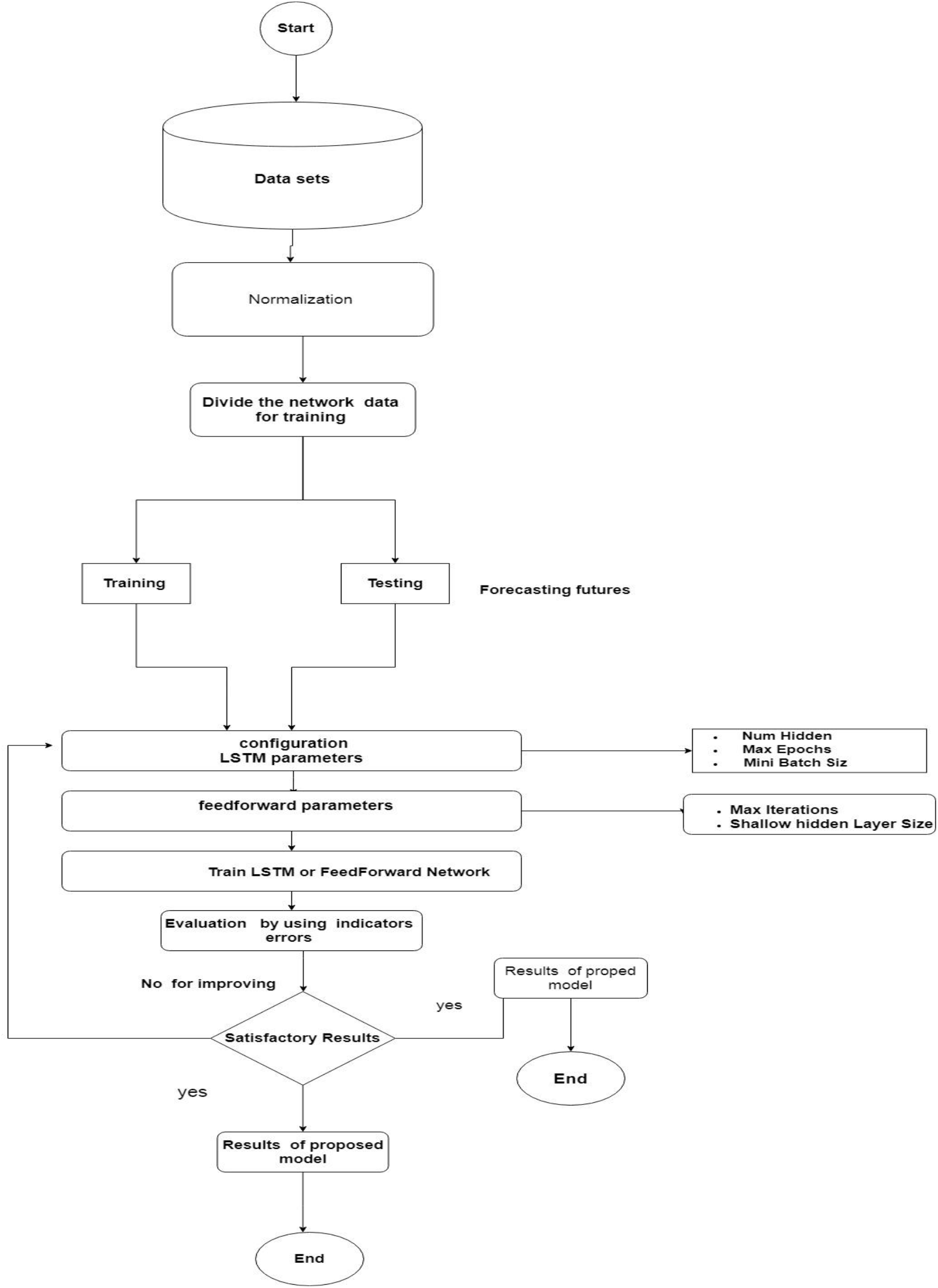
Flow steps of LSTM model.

#### 2.3.2 Holt-Trend model

Exponential Smoothing models are one of important prediction approach widely used in industry and commerce. Exponential smoothing method is the generalization of moving average technique. Exponential Smoothing models are one of prediction approaches that use the stationary time series data. The idea behind the Exponential Smoothing is to smooth the original time series data for forecasting future values.

Holt-Trend Exponential Smoothing model (HTES) is similar to Weighted Exponential Smoothing. However, it uses a trend estimator that changes over time. Furthermore, when the parameters *β*_0_ and *β*_1_ are slowly changing over time. Holt-Trend Exponential Smoothing method can be employed to the time series observations when neither _0 or 1 is changing over time, regression can be used to forecast future values of *y_t_*. Holt-Trend Exponential smoothing method approach has two smoothing constants, denoted by *α* and *β*. There are two estimators *ℓ*_*t*−1_ and *b*_*t*−1_; the *t* − 1 refers to the estimate of the level of the time series constructed in time period *t* − 1 (this is typically called the level component). The *b*_*t*−1_ refers to the estimate of the growth rate of the time series constructed in time period *b*_*t*−1_ (this is typically called the trend component). For fitting the holt-trend algorithm use *ℓ*_0_ and *b*_0_. Figure 8 demonstrates the Holt-Trend algorithm steps to predict the COVID-19 and *P*_*i*_ is prediction output, *f*_*i*_ is forecasting future values. Table 3 shows the important parameters that are fitted in the Holt-Tend model.

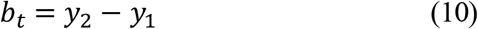

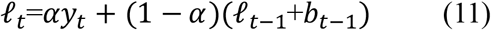

**Table 3.** Shows parameters of Holt-Trend algorithm.

| Parameters of Holt-Trend algorithm |  |
| --- | --- |
| Alpha | 0.1, 0.5, 0.15 |
| Beta | 0.20 |

**Figure 8.**
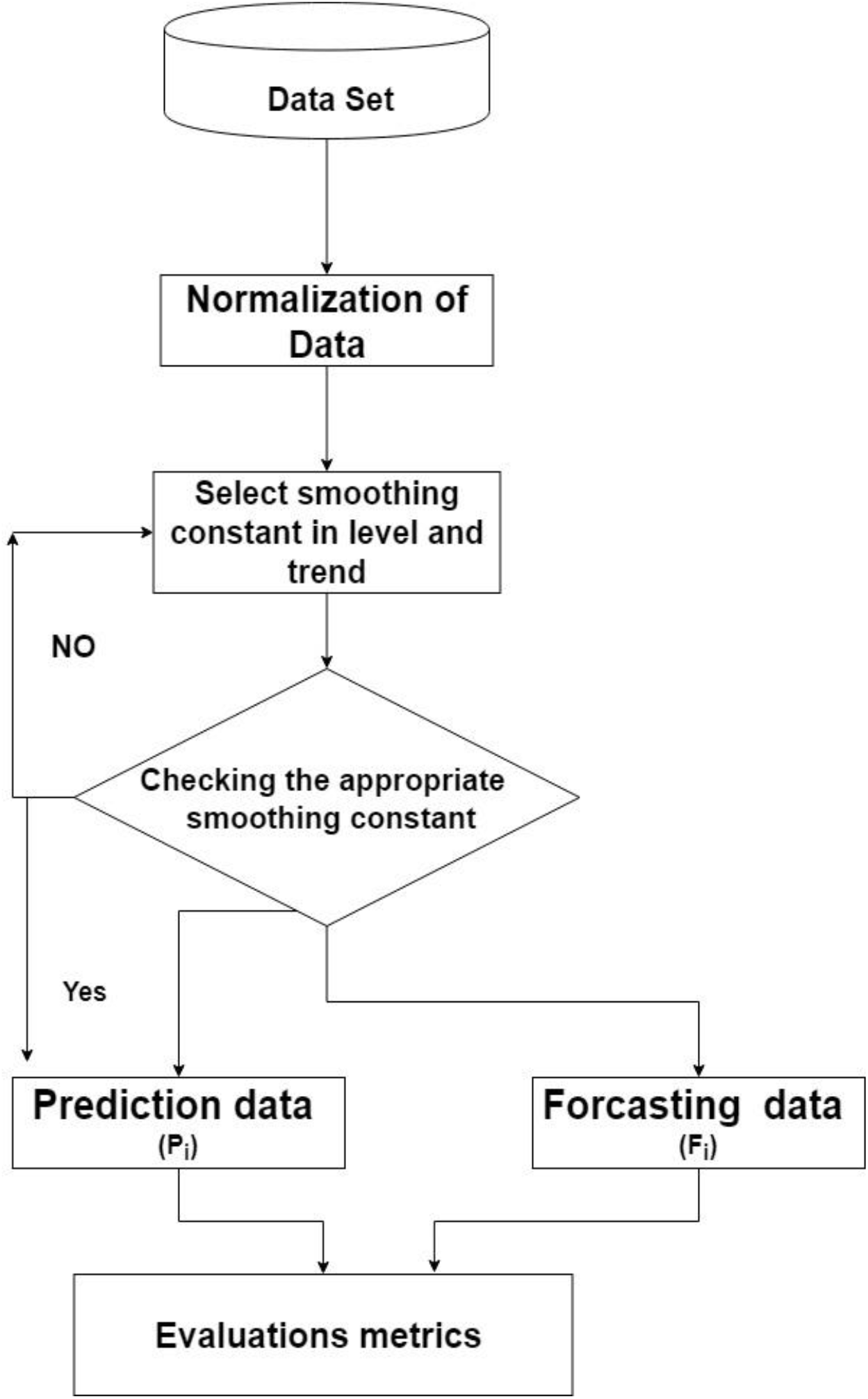
Flow steps of Hot-Trend model.

Where *b_t_* and *ℓ_t_* is trend level used to estimate the trend

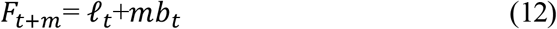

Where (*m* = 1,2,3…) *and F*_*t+m*_ is forecast future value

### 2.4. Model Evaluation Criteria

To evaluate the performance of the LSTM and Holt-Trend model, MSE, RMSE and Mean error metrics are applied to evaluate the proposed models. These standard metrics have the capability to measure the LSTM and Holt-Trend models by finding out the prediction errors.

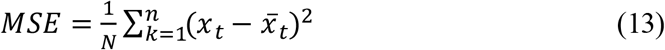

Where, *x_t_* is observed responses, 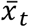 are estimated responses and *N* is the total number of observations.

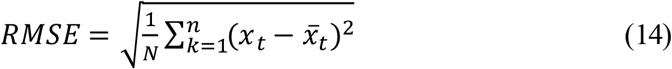

Where, *x_t_* is observed responses, 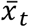 are estimated responses and *N* is the total number of observations.

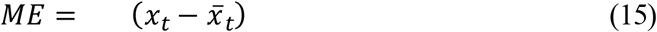

Where, *x_t_* is observed responses, 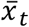 are estimated responses.

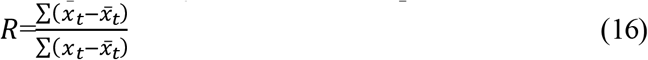

## 3. Results

Our analyses used data of 55 days that were sourced from WHO in a period of time from Jan 21^th^, 2020 to April 15^th^ 2020. The Min-Max method was applied for normalization purposes. Three countries namely Saudi Arabia, Spain and Italy are considered to test and evaluate the proposed model for prediction COVID-19. Two advance time series models are applied to predict the confirmed and death cases of COVID-19. Two experiments are conducted to obtain the prediction result. These two experiments are presented in subsection:

### 3.1 Results analysis of the LSTM model

In this section, we evaluate the performance of the LSTM deep learning approach. In this experiment, the deep learning algorithm was proposed, the real data set have been divided into 80% training and 20% testing. The training data is considered as self-similar prediction whereas the testing is forecasting and validation of the proposed model. To assess the performance of the prediction model, the evaluation metrics namely MSE, RMSE, ME and R values were employed to examine and evaluate the LSTM model. The LSTM is used to predict the confirmed and death cases.

Table 4 summarises the empirical results of the LSTM to predict confirmed cases of COVID-19. Three countries are considered in this proposed study to test the LSTM to predict the COVID-19. The results of the LSTM model used to predict the confirmed cases in Saudi Arabia are 000132, 0.0363, 0.00023, 0.0370 with respect to MSE, RMSE, mean error and standard deviation error respectively in training data whereas as the testing are MSE = 0.8111,RMSE = 0.9006, Mean error = 0.8811 and St.D = 0.228.

**Table 4.** Results of LSTM to predict confirmed cases of COVID-19.

| Training data |  |  |  |  |  |
| --- | --- | --- | --- | --- | --- |
|  | MSE | RMSE | Mean Error | St.D | R |
| Saudi Arabia | 0.00132 | 0.0363 | 0.00023 | 0.0370 | 99.74 |
| United State | 0.0885 | 0.297 | 0.00023 | 0.3014 |  |
| Italy | 0.0028 | 0.01676 | 2.0558e-05 | 0.0169 | 99.94 |
| Spain | 0.00020 | 0.01426 | 3.5212e-05 | 0.01441 | 99.91 |
| Testing data |  |  |  |  |  |
|  | MSE | RMSE | Mean Error | St.D | R (%) |
| Saudi Arabia | 0.8111 | 0.9006 | 0.8811 | 0.228 | 99.86 |
| United State | 0.9907 | 0.995 | 0.9130 | 0.433 |  |
| Italy | 0.1426 | 0.3777 | 0.3585 | 0.1258 | 98.86 |
| Spain | 0.0527 | 0.2296 | 0.1949 | 0.1288 | 99.16 |

As shown in Table 4, the results obtained from the LSTM model to predict confirmed cases in Italy, in the training data the prediction results are 0.0028, 0.01676, 2.0558e-05 and 0.0169 in terms of MSE, RMSE, mean error and standard deviation error respectively. It is noted that the performance of the LSTM is higher and the prediction errors are very high compared with the LSTM of Saud Arabia data. For validation and testing the proposed model we have divided 20 testing and the results of the LSTM in the testing validation are MSE = 0.1426, RMSE = 0.377, Mean error = 0.3585 and St.D 0.1288. Similarly, the LSTM model has been implemented on the Spain data set. The performance of the LSTM to predict the confirmed case in Spain, in the training 0.00020, 0.01426, 3.5212e-05 and 0.01441 according to MSE, RMSE, mean error and standard deviation error respectively, the testing MSE = 0.0527, RMSE = 0.2296, Mean error = 0.1949 and St.D 0.1288. Figure 9 (a, b and c) displays the time series plot of the training data for the three countries. The blue line refers to the observation data and the red line shows the prediction output. It is noted that the prediction data is very close to the observation data, this approved that LSTM is an appropriate model for the prediction of the COVID-19. The prediction is very less. It indicates that the LSTM model is more efficient and effective. To handle the COVID-19 and find out the numbers of confirmed cases that will be discovered in future.

**Figure 9.**
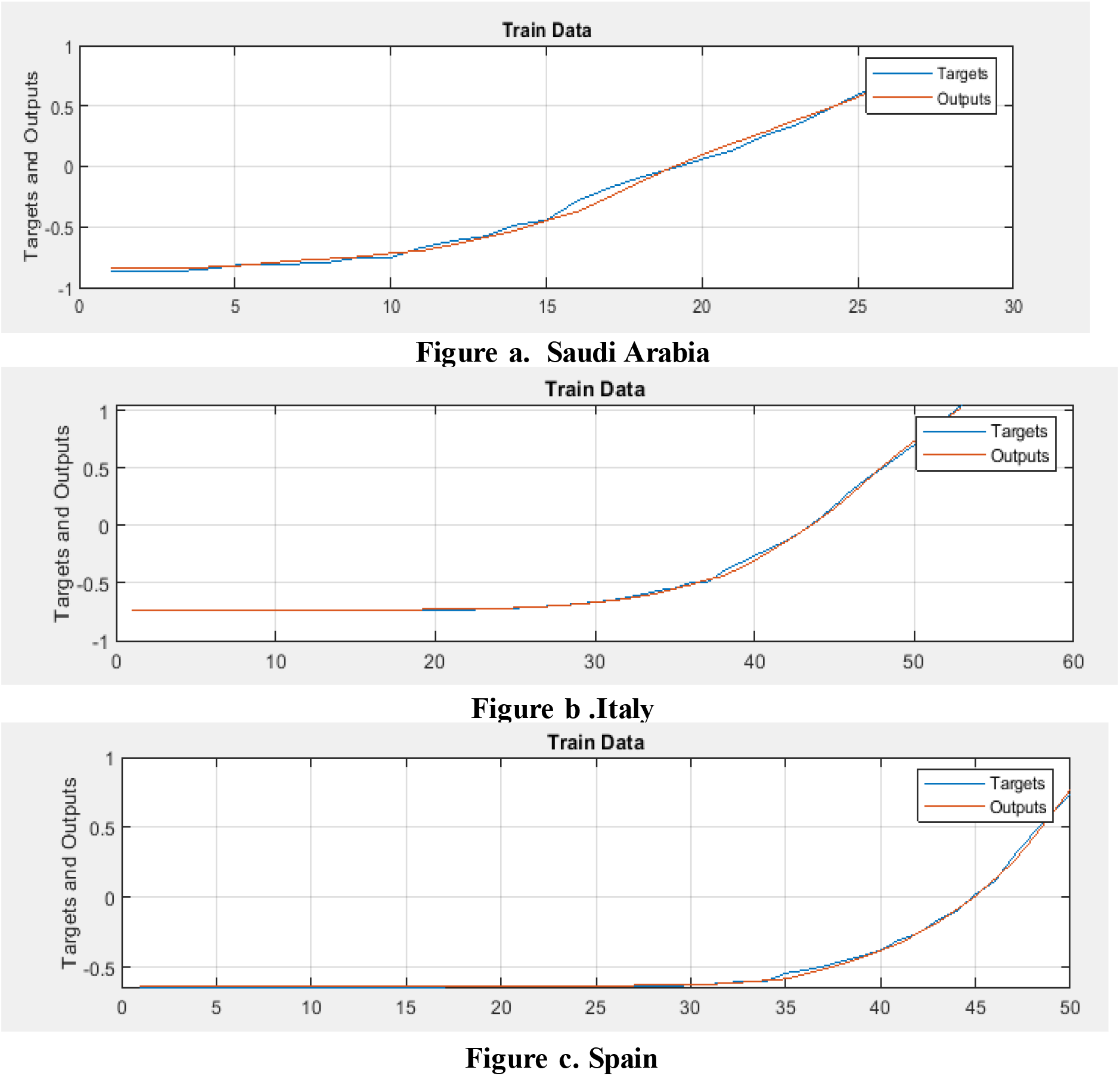
The performance of LSTM model to predict numbers of confirmed cases in a) Saudi Arabia, b) Italy and c) Spain.

Figure 10 (a, b and c) provides information regarding the graphical performance of LSTM model for predicting the number of confirmed cases of COVID-19 in three countries namely Saudi Arabia, Italy and Spain. The graphically representation shows that the training and testing are fitting in the regression line which indicates that the proposed model was appropriate to predict the cases of COVID-19. The percentage value of R is very high between the observation value and output value. The empirical results of the validation process of the LSTM model also indicate that the constructed model can achieve equally impressive performance.

**Figure 10.**
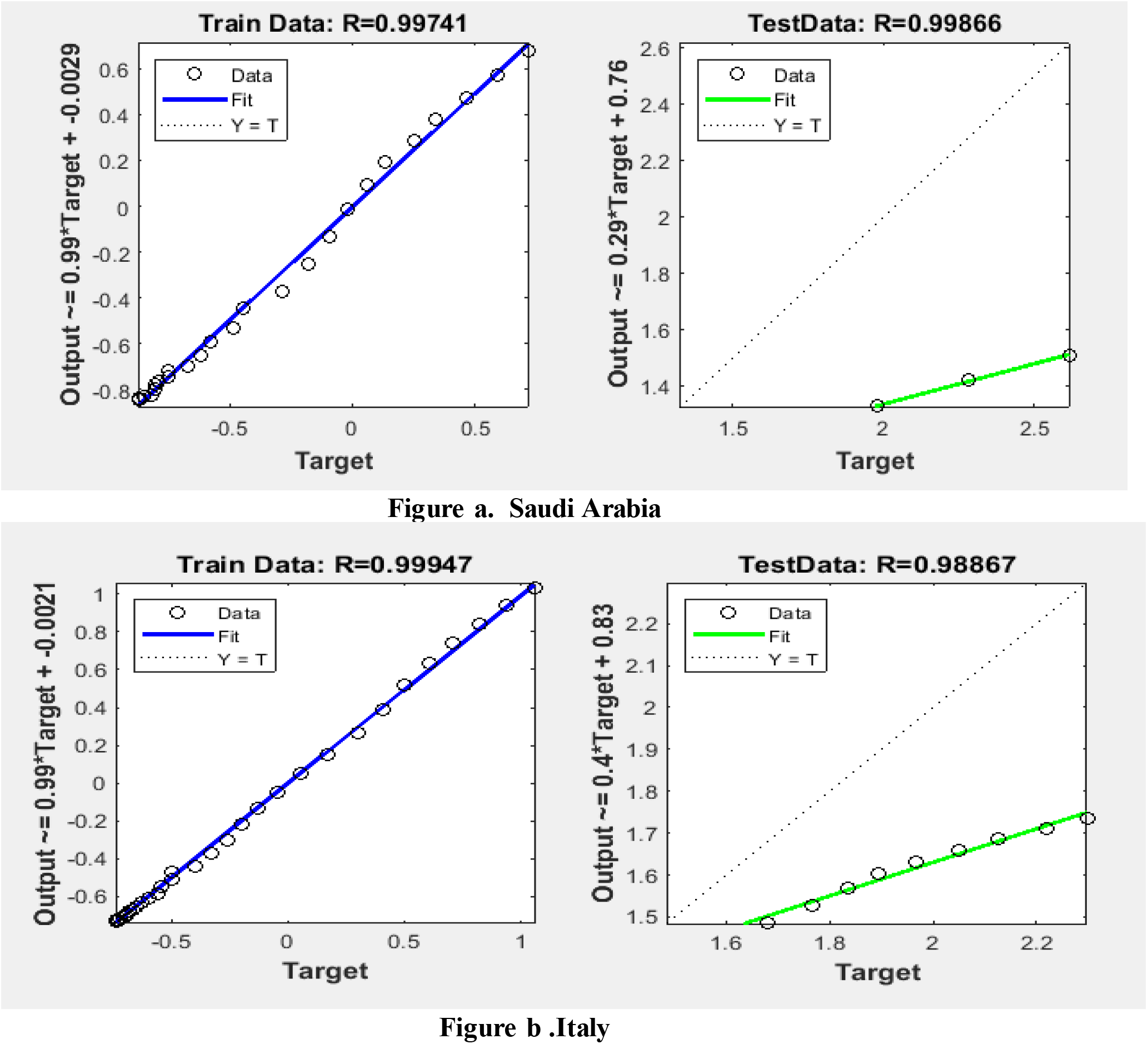
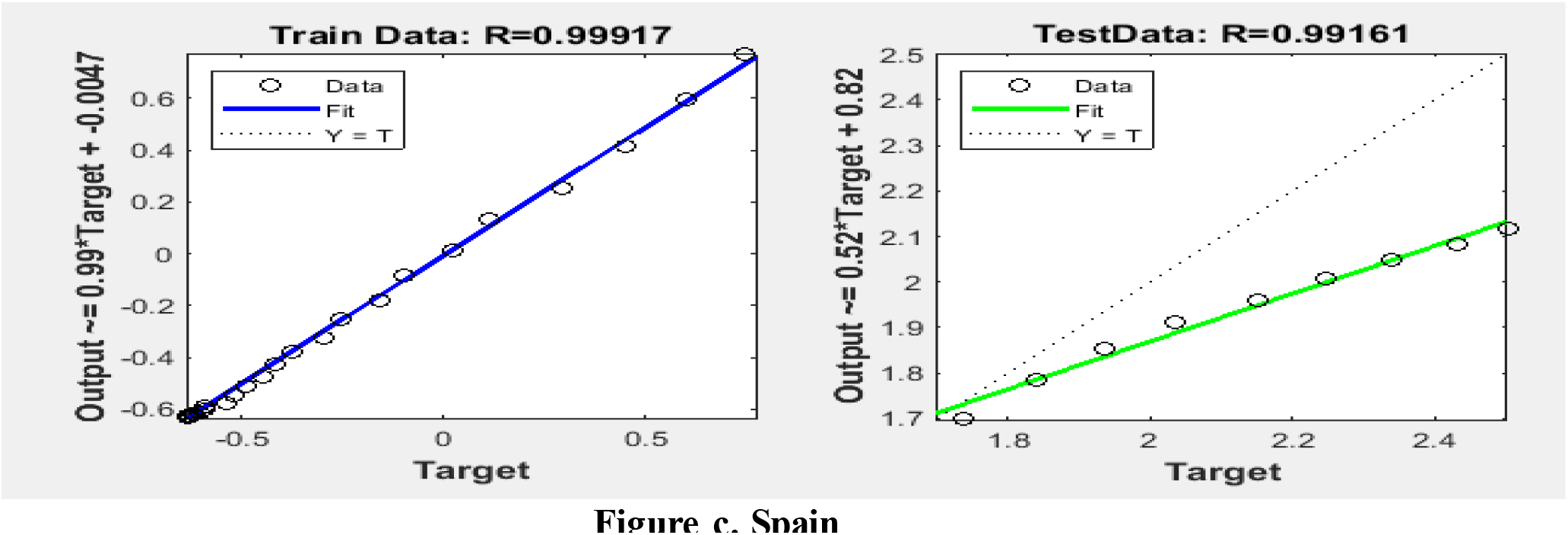
Scatter plot for flow in the validation confirmed cases in a) Saudi Arabia, b) Italy and c) Spain.

In this subsection, we focus on predicting the numbers of death in these countries. Table 5 illustrates prediction results of the LSTM for numbers of death cases in three countries namely Saudi Arabia, Italy and Spain. The LSTM has been applied to predict the number of deaths in these countries for discovering the number of deaths in future. For validating the proposed model, we have divided the data into training and testing whereas the testing considered forecasting the future values.

**Table 5.**
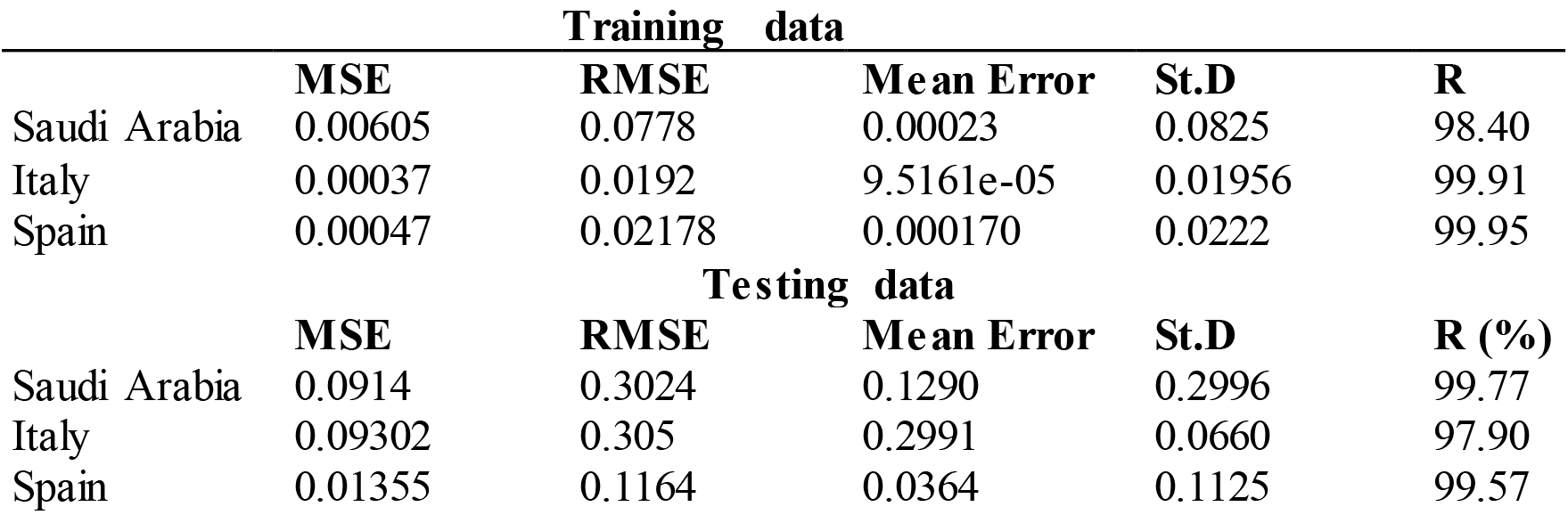
Results of LSTM to predict death cases of COVID-19.

The obtained prediction results of the LSTM model to predict the death cases in Saudi Arabia are 0.00605, 0.0778, 0.00023 and 0.082 with respect to MSE, RMSE, mean error and standard deviation error respectively in training and the performance of the proposed for forecasting the numbers of death in future are MSE = 0.0914, RMSE = 0.3024, Mean error = 0.2996 and St.D = 0.228. The similar accuracy of the proposed model to predict the numbers of death in the Italy country are MSE = 0.00037, RMSE = 0.0192, Mean error = 9.5161e-05 and St.D = 0.01956 in the training data, however, the results of proposed model in testing data are 0.09302, 0.305, 0.2991 and 0.0660 with respect to MSE, RMSE, mean error and standard deviation error respectively. The prediction results of the LSTM to predict the number of deaths in Spain are 0.00047, 0.02178, 0.000170 and 0.0222 according the standard metrics MSE, RMSE, mean error and standard deviation error correspondingly. In the results of testing data are 0.01355, 0.1164, 0.0364 and 0.1125 in terms of the standard metrics. Figure 11 (a, b and c) shows the time series plot of training data to predict number of deaths in three countries. The blue colour represents the observation data, the red colour represent the prediction value. It is noted that the prediction line very adjacent to the actual data.

**Figure 11.**
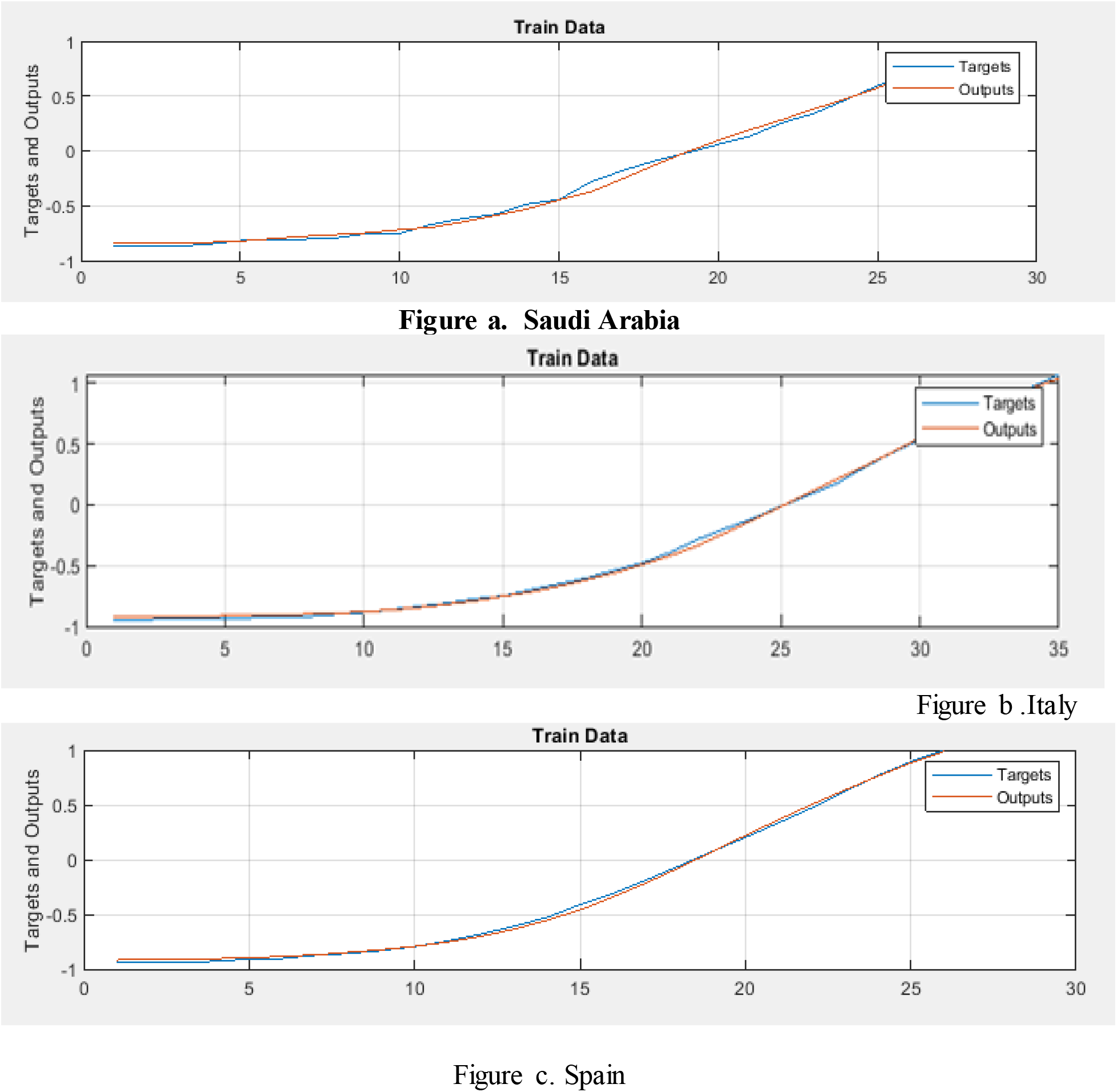
The performance of LSTM model to predict numbers of death cases in a) Saudi Arabia, b) Italy and c) Spain.

Figure 12 (a, b and c) illustrates regression plots to discover the correlation between the observation data of number of death cases and prediction output. The graphically representation shows that the training and testing data are fitting in the regression line, indicates that the proposed model was appropriate to predict the number of death cases of COVID-19. The percentage value of correlation metrics are very high between the observation of number of death value and output value. The empirical results of the validation process of the LSTM model also indicate that the constructed model can achieve equally impressive performance even if some parameters such as the number of units or the number of input variables have been changed.

**Figure 12.**
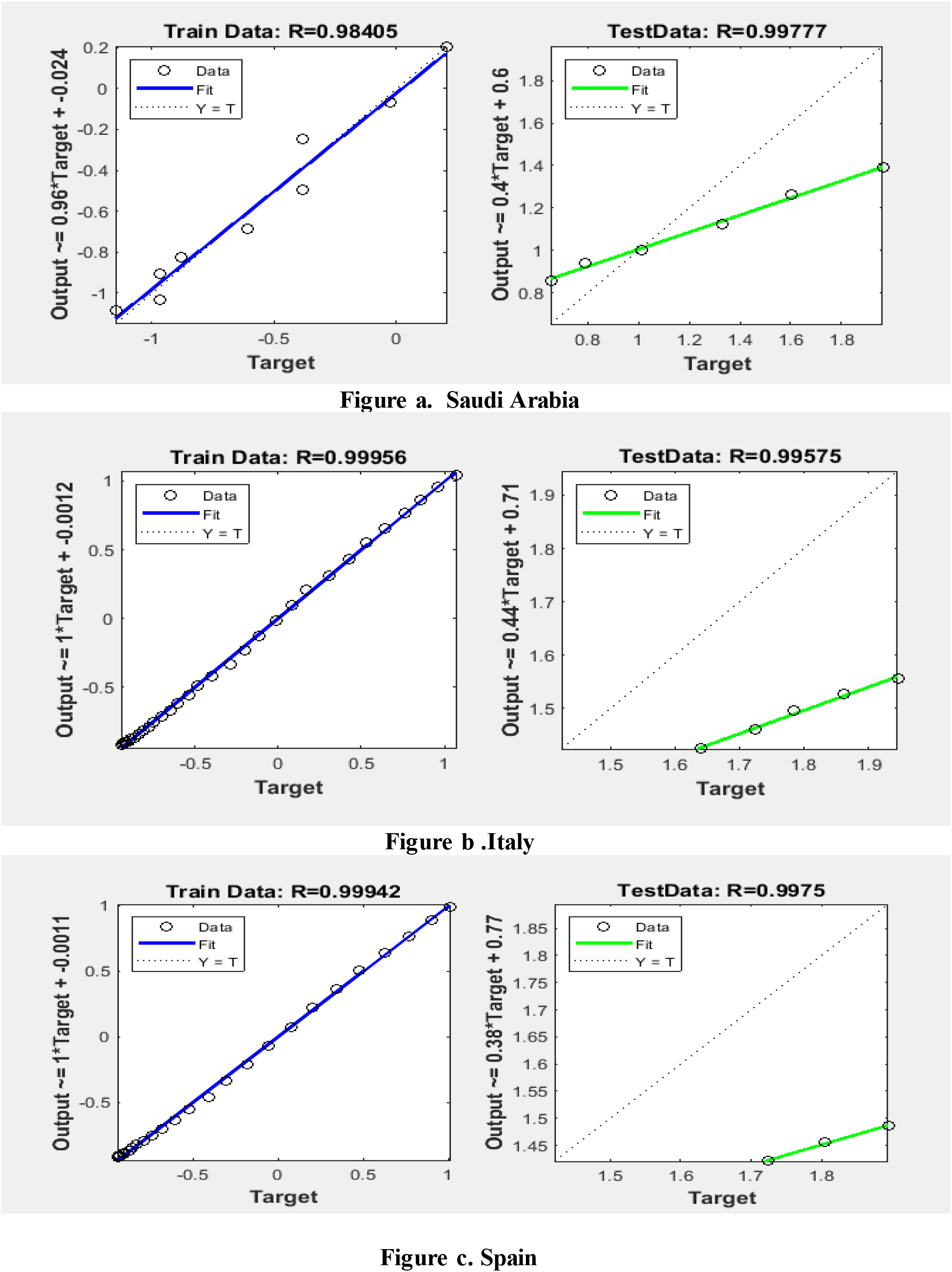
Scatter plot for flow in the validation death cases in a) Saudi Arabia, b) Italy and c) Spain.

### 4.2. Results analysis of the Holt-Trend model

The Holt-Trend model is one type of Exponential Smoothing models, it is used to predict the trend data. The Holt-Trend model has two smoothing constants at level and trend. In this experiments we have taken different parameters at level and trend for obtaining higher prediction, at level there different parameters values are considered alpha 0.1, 0.5 and 0.15 whereas the trend values are some beta = 0.20. MSE metric has been used to measure the best parameters, it is noted that the level 0.15 and trend 0.20 are appropriate because the prediction errors are very less according to the standard evaluation metrics.

Table 6 shows that the result of the Holt-trend model for predicting the numbers of confirmed cases in three different countries. The prediction results of Hot-Trend model to predict the numbers of confirmed case in the Saudi Arabia are 0.007, 0.085, 0.0377 with respect to MSE, RMSE, mean error and standard deviation error respectively. Similarly the analysis result for prediction confirmed case in Italy are MSE = 3.8378e-04, RMSE = 0.0196 Mean Error = 0.0082, it is noted that the prediction errors are very less. The Holt-Trend model is applied to predict the numbers confirmed cases in Spain, the empirical results are 5.1711e-04, 0.0227 and 0.0085 according the standard metrics MSE, RMSE and Mean error. Figure 13 (a, b and c) displays performance of the Holt-Trend for predicting the number of confirmed cases of COVID-19 in three countries, it is indicated that the observation data very close to the output of prediction data. From the graphics, the green line is observation data and red, blue and white are prediction output obtained from three different parameters’ values. It is observed that the parameters’ values of level = 0.15 and trend 0.20 are more suitable to the prediction of the number of confirmed cases of COVID-19.

**Table 6.** Results of Holt-Trend model to predict confirmed cases of COVID-19.

|  | MSE | RMSE | Mean Error | R |
| --- | --- | --- | --- | --- |
| **Saudi Arabia** | 0.0074 | 0.085 | 0.0377 | 99.06 |
| **Italy** | 3.8378e-04 | 0.0196 | 0.0082 | 99.96 |
| **Spain** | 5.1711e-04 | 0.0227 | 0.0085 | 99.94 |

**Figure 13.**
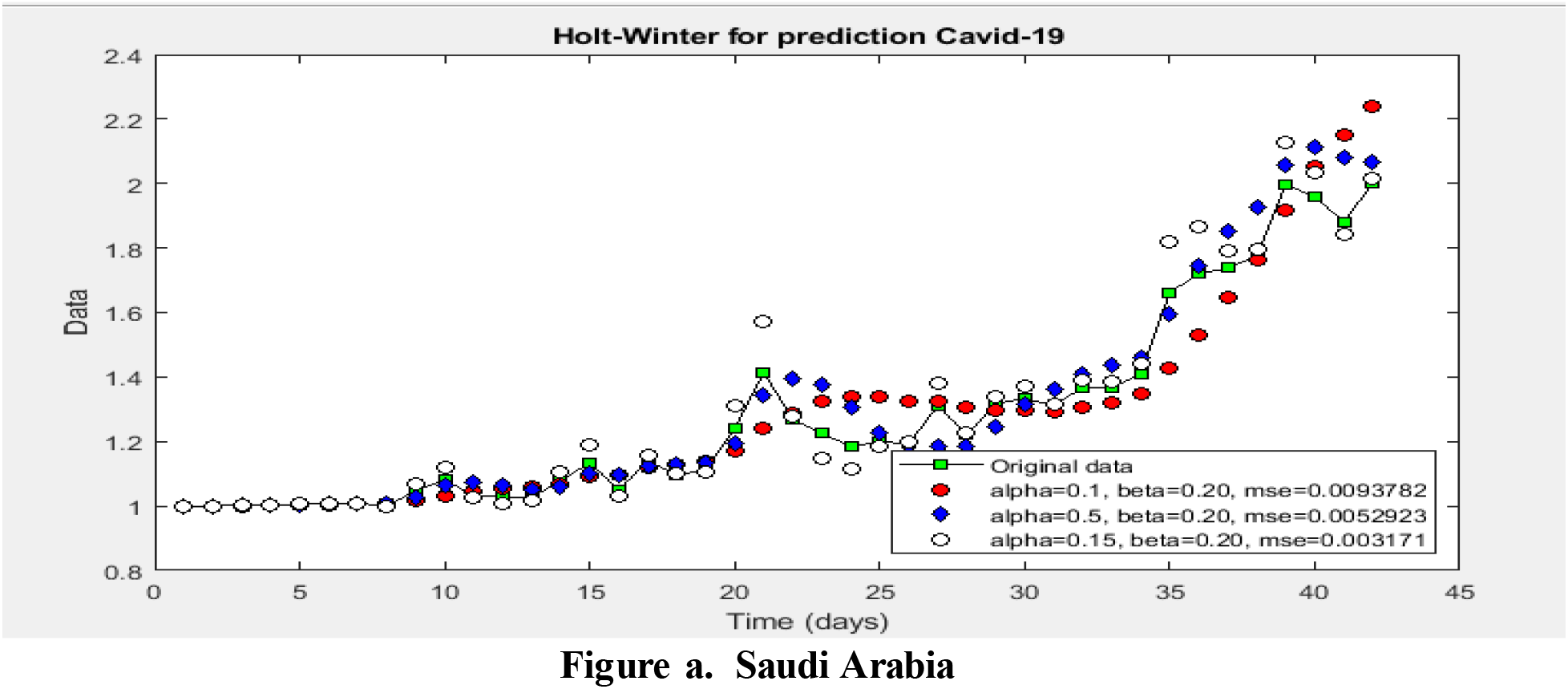
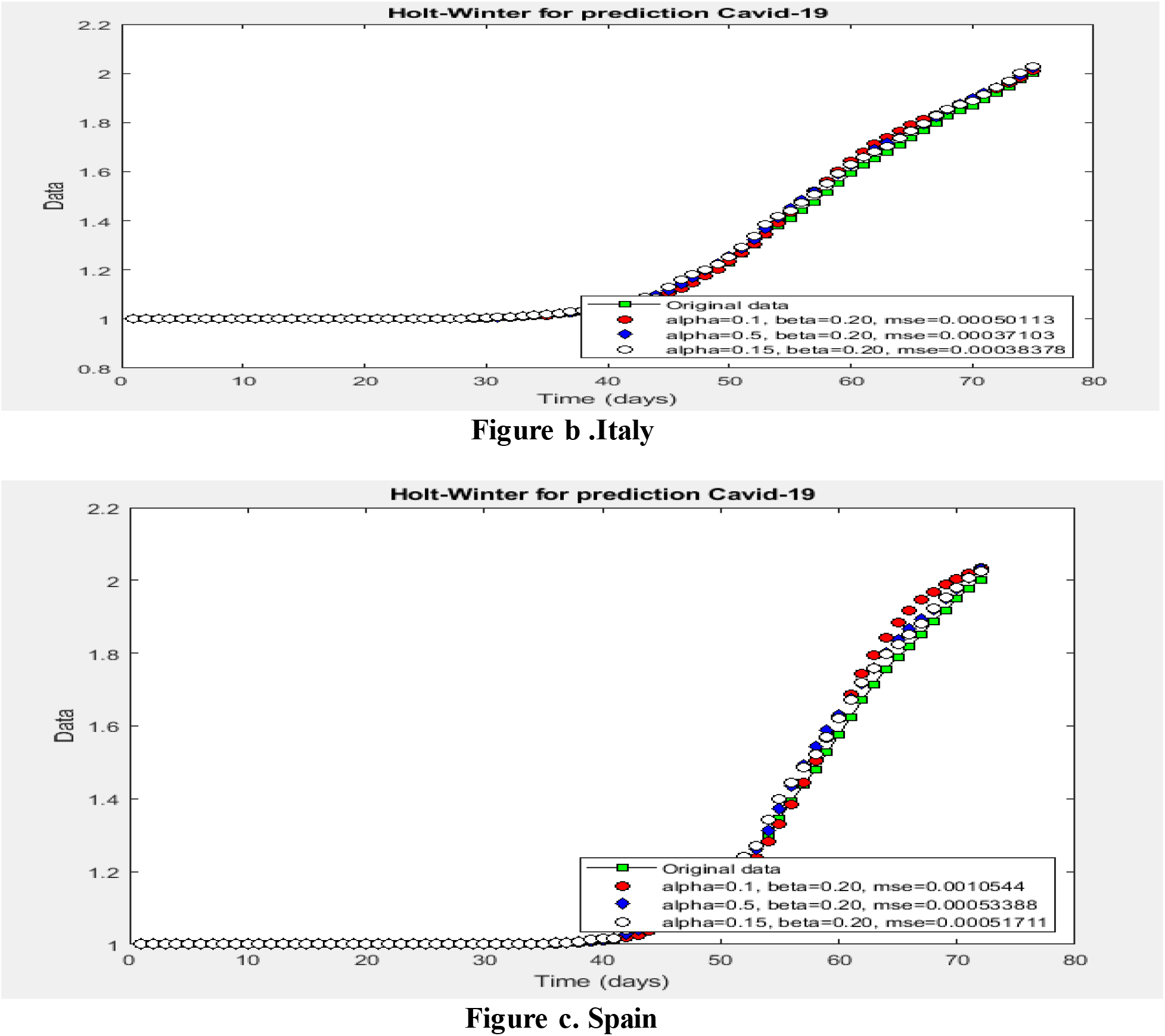
The performance of Holt-Trend model to predict numbers of confirmed cases in a) Saudi Arabia, b) Italy and c) Spain.

Figures 14 (a, b and c) shows the performance of the Hot-Trend to the number of confirmed cases in three countries by using regression plot. The graphically representation is shown the percentage correlation between the observation and prediction output. It is noted that the correlation between the observation numbers of confirmed cases in Saudi Arabia and prediction out is 99.64% whereas the correlation percentage for prediction number of confirmed cases in Italy is 99.96%. Correspondingly the regression plot is shown in Spain is 99.94%. Overall, the Holt-Trend has the ability to predict the numbers of the confirmed cases of COVID-19 with higher percentage.

**Figure 14.**
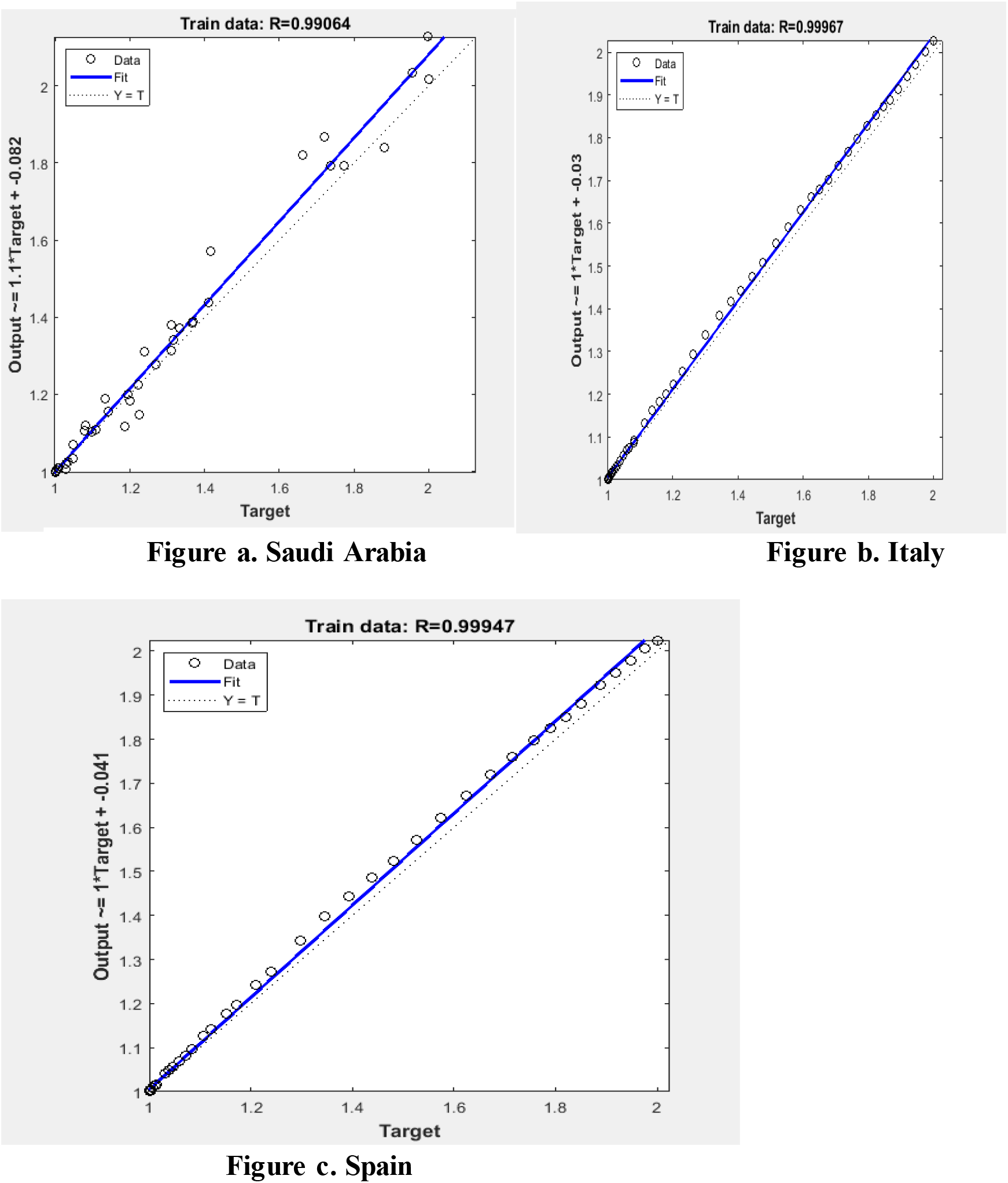
Scatter plot for flow in the validation numbers of confirmed cases in a) Saudi Arabia, b) Italy and c) Spain.

In this section, the focus is on forecasting the future values of numbers of confirmed cases in theree countries. We have forcasted future valus in time interval one month from 15-4-2020 into 15-05-2020. The Holt-Trend model is applaied to forcat the future values by using obsrevatiuon data that are collected from WHO. Figures demonstrate the performance of the Holt-Trend model to forecast the future values, it is observed that the trend growing up, this indicates that the numbers of confirmed cases will increase. Figure **15 (a, b and c)** illustrates graphically the prediction performance of Holt-Trend model to forecast the number of confirmed cases in future. It is indicated that the trend growing up, this indicates that the number of the confiremd casses will increase.

**Figure 15.**
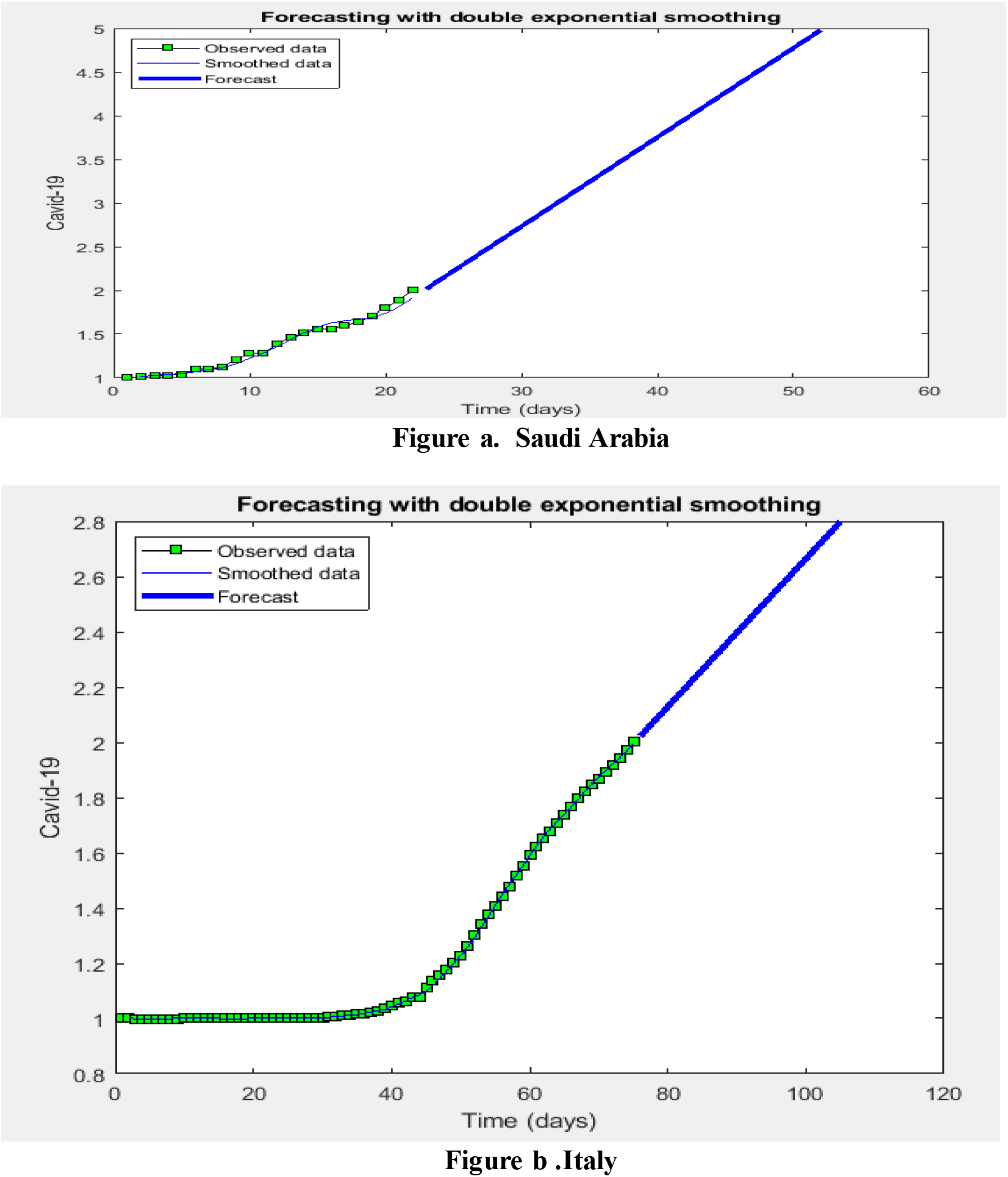
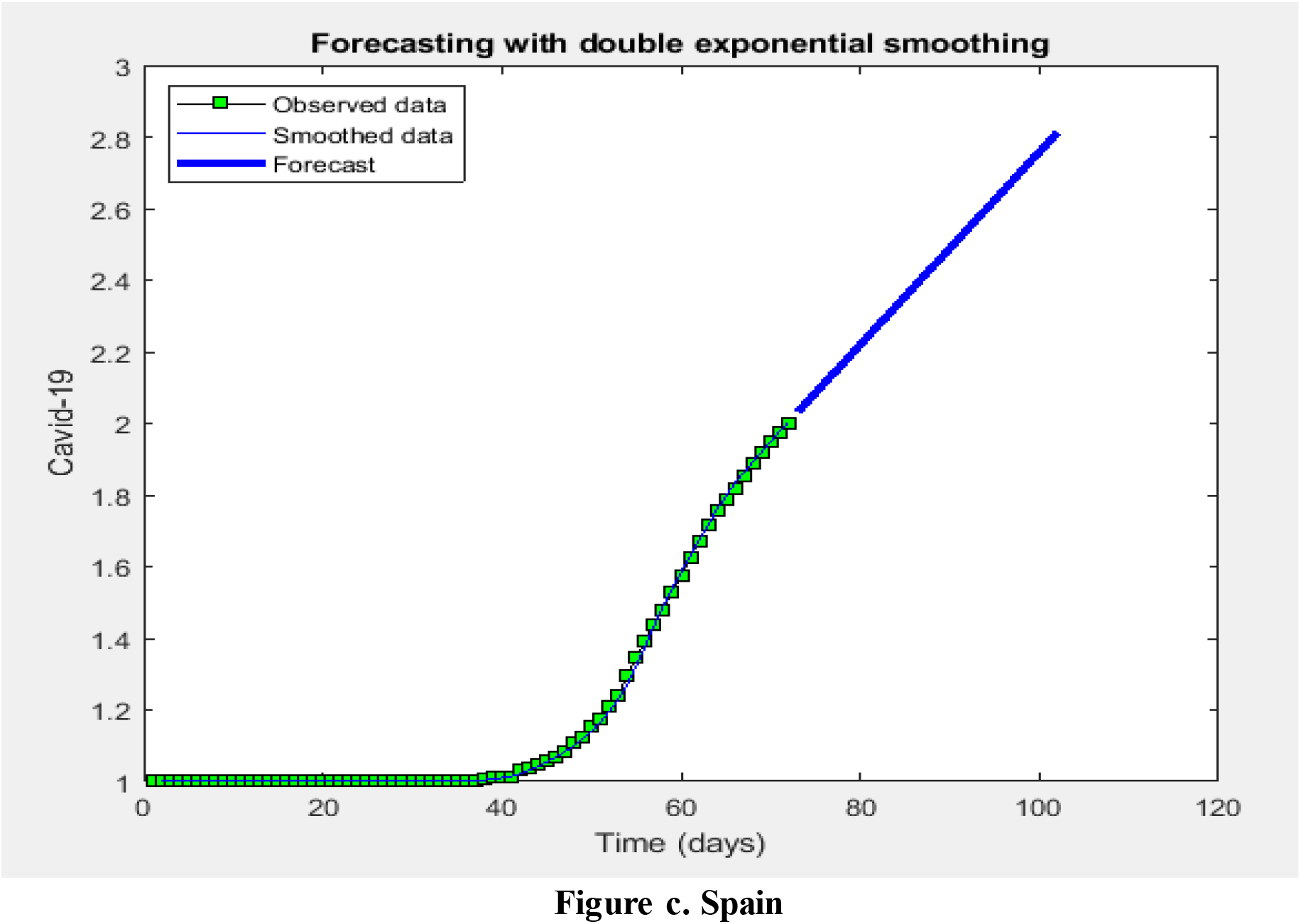
Performance of Holt-Trend to forecast the numbers of confirmed in future for a) Saudi Arabia, b) Italy and c) Spain.

In this section, the Hot-Trend model has been applied to predict the number of deaths in three countries. The Holt-Trend model depends on two constant values at level and trend. It is noted that the level 0.15 and trend 0.20 is appropriate because the prediction errors are very less according the standard evaluation metrics.

Table 7 shows the results of the Holt-trend model for predicting the number of death cases in three different countries. The prediction results of Hot-Trend model to predict the numbers of death cases in the Saudi Arabia are 0.0030, 0.0546, 0.0248 with respect to MSE, RMSE, mean error and standard deviation error respectively. Similarly, the analysis result for prediction death case in Italy country are MSE = 5.6995e-04, RMSE = 0.0239 Mean Error = 0.0082, it is noted that the prediction errors very less. The Holt-Trend model is applied to predict the numbers of confirmed cases in Spain country, the empirical result are 9.3129e-04, 0.0304 and 0.0145 according the standard metrics MSE, RMSE and Mean error. Figure 16 (a, b and c) displays the performance of the Holt-Trend for prediction of the number of death cases of COVID-19 in three countries, it is indicated that the observation data is very close to the output of prediction data. From the graphics, the green line is the observation data and red, blue and white are prediction output obtained from three different parameters’ values. It is observed that the parameters’ values of level = 0.15 and trend 0.20 are more suitable for the prediction of the number of death cases in COVID-19.

**Table 7.** Results of Holt-Trend to predict death cases of COVID-19.

|  | MSE | RMSE | Mean Error | R |
| --- | --- | --- | --- | --- |
| **Saudi Arabia** | 0.0030 | 0.0546 | 0.0248 | 99.80 |
| **Italy** | 5.6995e-04 | 0.0239 | 0.0117 | 99.94 |
| **Spain** | 9.3129e-04 | 0.0304 | 0.0145 | 99.94 |

**Figure 16.**
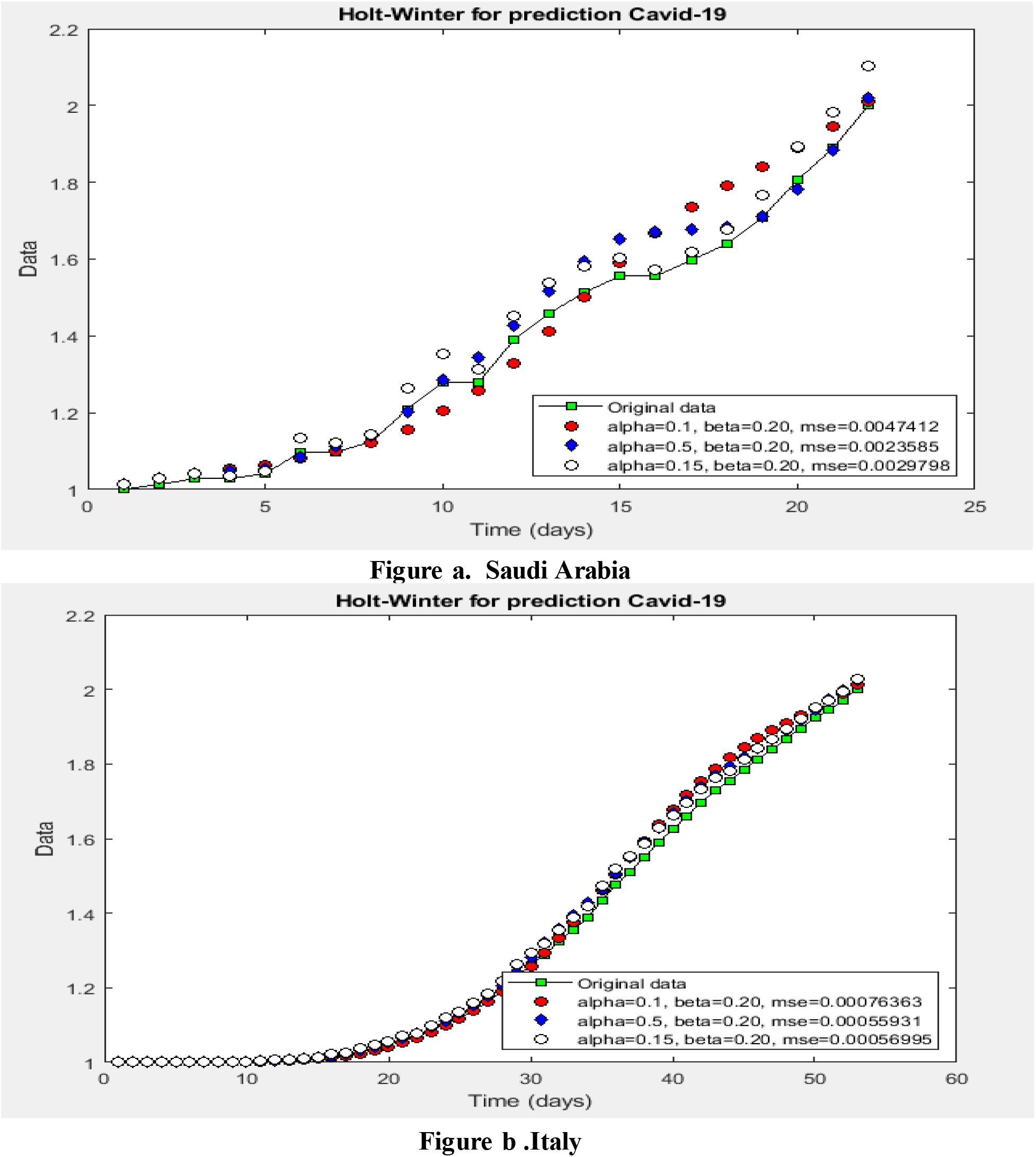
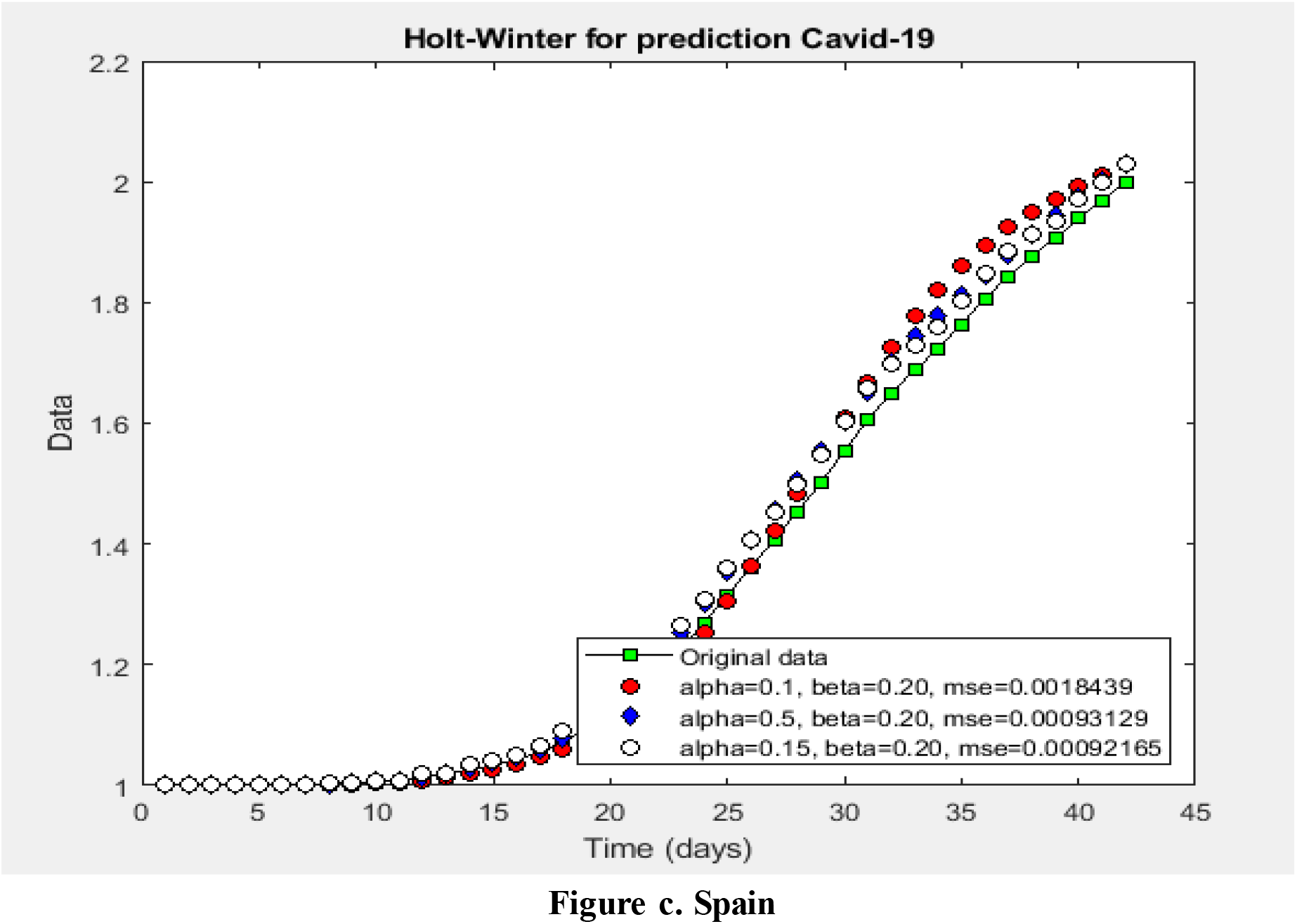
The performance of Holt-Trend model to predict numbers of death cases in a) Saudi Arabia, b) Italy and c) Spain.

Figure 17 (a, b and c) shows Scatter plots of correlation and time series analysis which were applied to assess the association between the observation data and the prediction output. Cross-correlation results are obtained as product-moment correlations between the observation data and prediction output. The time dependence between two variables is termed as lag, the Lag values indicate the degree and direction of associations between the observation values and prediction values. It is noted that the correlation between the observation numbers of death cases in Saudi Arabia and prediction is 99.80% whereas the correlation percentage for predicting the number of death cases in Italy is 99.96%. Correspondingly the regression plot in Spain is 99.93%. Overall, the Holt-Trend has the ability to predict the number of death cases of COVID-19 with higher percentage.

**Figure 17.**
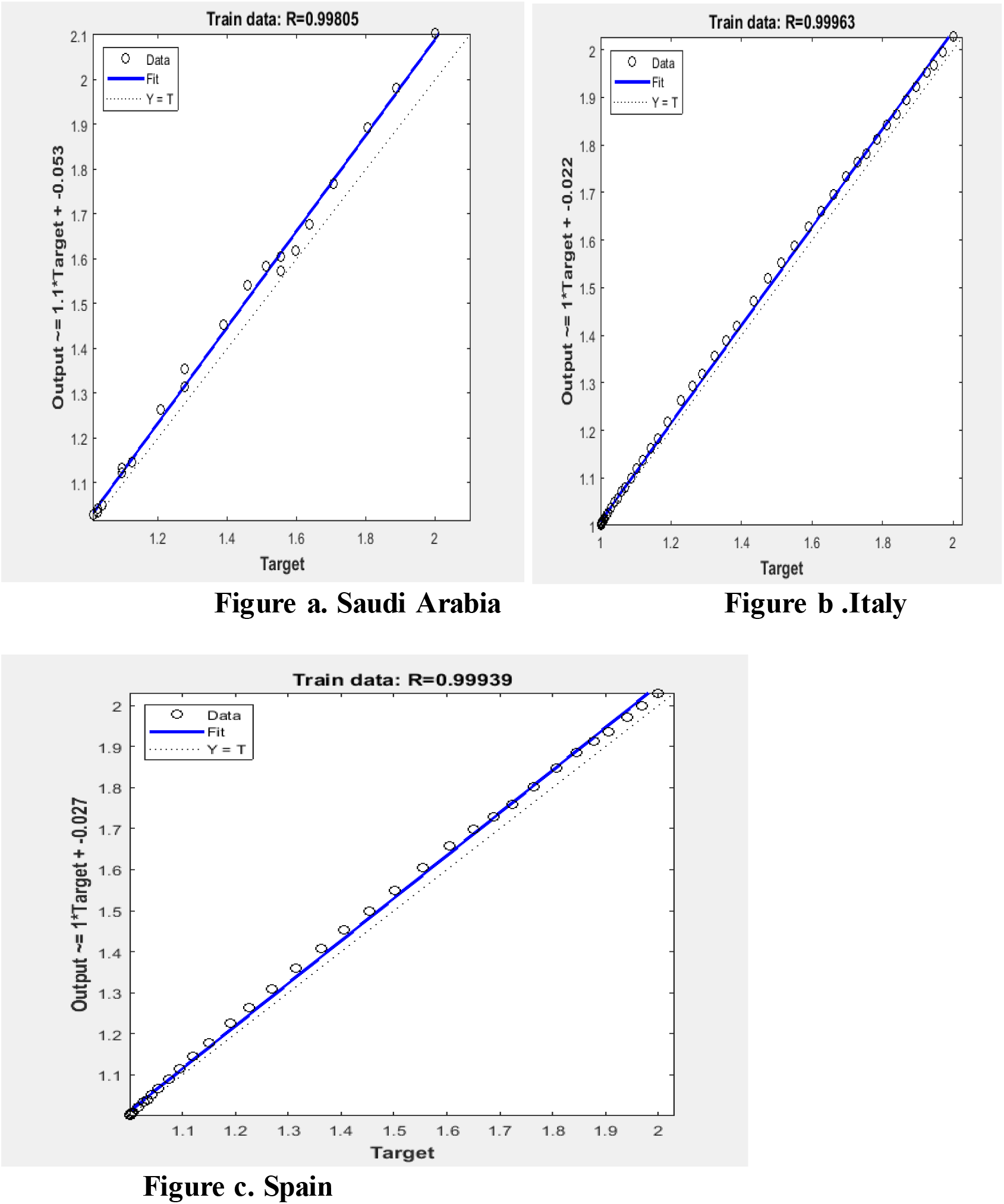
Scatter plot for flow in the validation of death cases in a) Saudi Arabia, b) Italy and c) Spain.

In this section, we focus on forecasting the future values of the number of death cases in theree countries. We have forcasted future values in time interval of one month from 15-4-2020 into 15-05-2020. The Holt-Trend model is pplaied to forcast the future values by using obsrevatiuon data that are collected from WHO. Figures demonstrate the performance of the Holt-Trend to forecast the future values, it is observed that the trend growing up, this indicates that the number of confirmed cases will increase. Figure 18 (a, b and c) illustrates graphically the prediction performance of Holt-Trend model to forecast the number of the confirmed cases in future. It is indicated that the trend is growing up, this suggests that the number of confiremd cases will increase.

**Figure 18.**
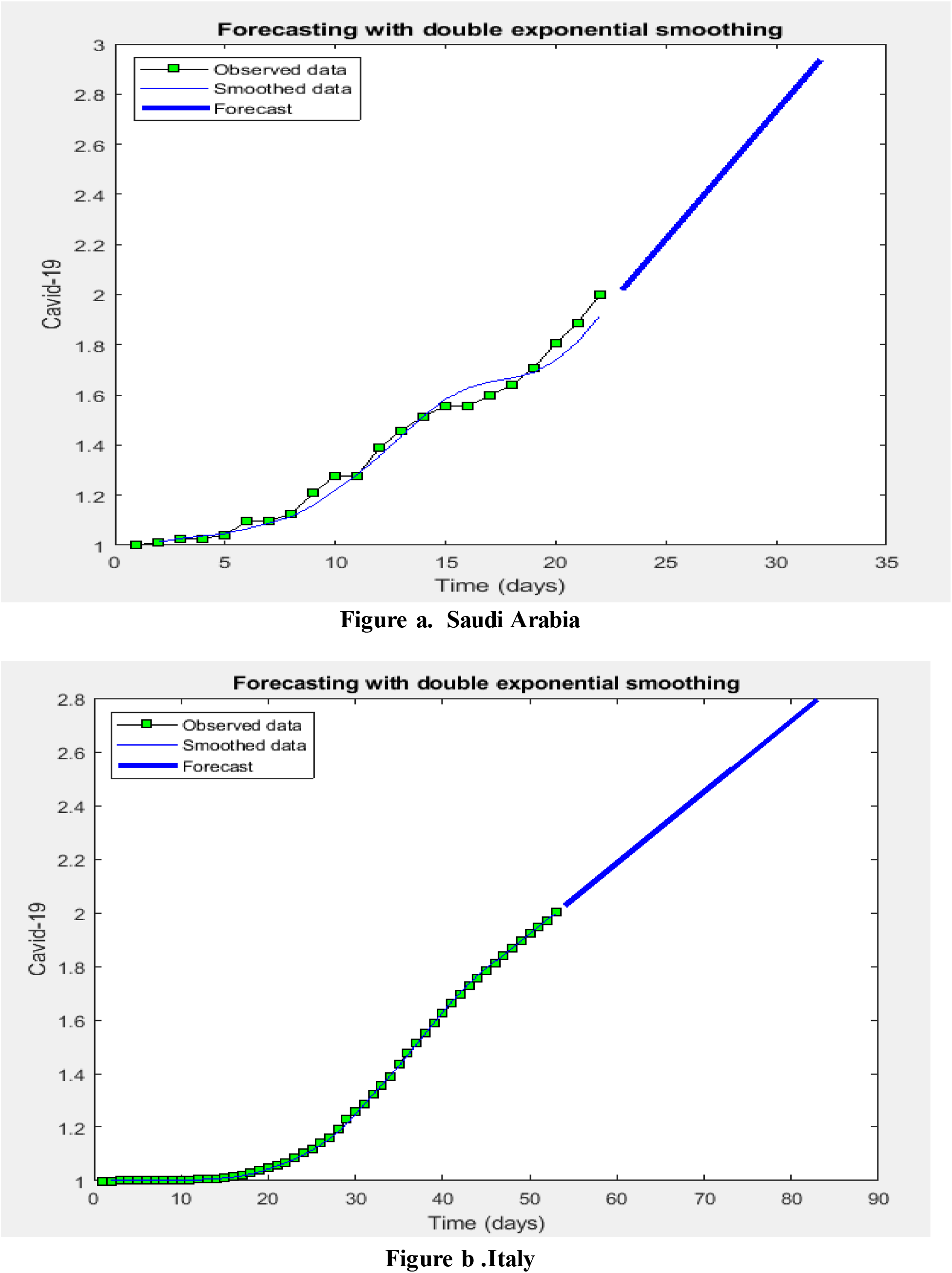
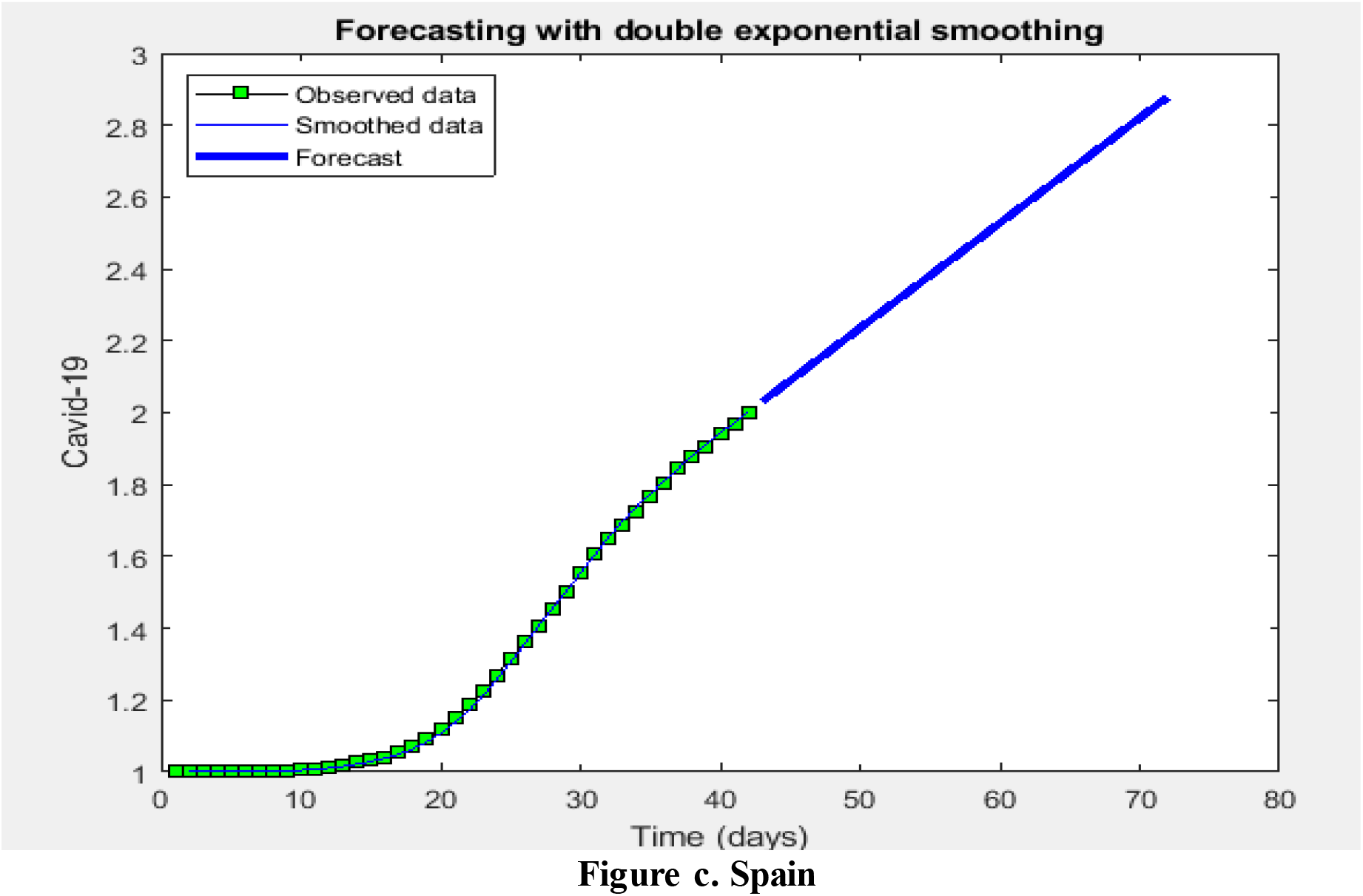
performance of Holt-Trend to forecast the numbers of death in future for a) Saudi Arabia, b) Italy and c) Spain.

## 4. Conclusion

This study applied deep learning and Holt-Trend model to predict the risk of COVID-19 outbreak based on the real time data collected from the WHO. The main object of the proposed model is to find out the number of the confirmed and death cases in future. The proposed models can be used to estimate the risk of COVID19 in future. The Max-Min normalization is applied to save the range of data. These algorithms were applied to predict numbers of confirmed and death cases. In the LSTM algorithm, the data have been divided into the 80% training used for self prediction whereas 20% testing for validation and future forecasting. The statistical Holt-trend was applied to predict the numbers of confirmed and death cases. The Holt-Trend has applied to all data to predict self prediction, for validation we have forecasted future value in time interval of 30 days. The prediction results demonstrate that the LSTM and Holt-trend models can be effectively employed for the prediction of COVID-19 outbreak by using real time data that are gathered from WHO. A comparative prediction results between LSTM, Holt-Trend models is presented. It is noted that the proposed models showed effective performance according to MSE, RMSE, Mean error, and correlate of increment performance measures. Respectively, it is also observed that the LSTM and Holt-trend models are more satisfying to predict the COVID-19 cases. In future work, we will use the Google search terms to predict the COVID-19 cases.

## Notes

### Competing Interest Statement

The authors have declared no competing interest.

### Author Declarations

Your manuscript 8894808 Deep Learning and Holt-Trend Algorithms for predicting COVID-19 pandemic submitted to Computational Intelligence and Neuroscience has now passed the initial screening and has been assigned to an Academic Editor. We noted that your manuscript is associated with the ongoing coronavirus outbreak. In accordance with our commitment to the statement coordinated by the Wellcome Trust we strongly encourage you to post your manuscript on a preprint server such as medRxiv or bioRxiv and make all your data available on an open repository such as dryad or figshare if you haven t done so already. In addition we will provide the WHO with a copy of your manuscript in the next 2 days. Please rest assured that we are trying to do everything within our power to have your manuscript assessed as quickly as possible. If you have any questions in the meantime please do not hesitate to contact me.

## References

1. https://www.who.int/health-topics/coronavirus

2. https://www.oecd.org/economic-outlook/

3. McCarthy, M.J. Internet monitoring of suicide risk in the population. J. A_ect. Disord. 2010, 122, 277–279.

4. Sueki, H. The e_ect of suicide-related internet use on users’ mental health: A longitudinal study. Crisis 2013, 34, 348–353.

5. Sueki, H.; Yonemoto, N.; Takeshima, T.; Inagaki, M. The impact of suicidality-related Internet use: A prospective large cohort study with young and middle-aged Internet users. PLoS ONE 2014, 9, e94841. [CrossRef] [PubMed]

6. Sueki, H. Does the volume of Internet searches using suicide-related search terms influence the suicide deathrate: Data from 2004 to 2009 in Japan. Psychiatry Clin. Neurosci. 2011, 65, 392–394.

7. . Gunn, J.F.; Lester, D. Using Google searches on the Internet to monitor suicidal behavior. J. A_ect. Disord. 2013, 148, 411–412.

8. Yang, A.C.; Tsai, S.J.; Huang, N.E.; Peng, C.K. Association of Internet search trends with suicide death I Taipei City, Taiwan, 2004–2009. J. A_ect. Disord. 2011, 132, 179–184.

9. Tran, U.S.; Andel, R.; Niederkrotenthaler, T.; Till, B.; Ajdacic-Gross, V.; Voracek, M. Low validity of Google Trends for behavioral forecasting of national suicide rates. PLoS ONE 2017, 12, e0183149. [CrossRef]

10. Kristoufek, L.; Moat, H.S.; Preis, T. Estimating suicide occurrence statistics using Google Trends. EPJ Data Sci. 2016, 5, 32.

11. Callison-Burch, V.; Guadagno, J.; Davis, A. Building a Safer Community with New Suicide Prevention Tools. Facebook Newsroom 2017. Available online: https://newsroom.fb.com/news/2017/03/building-a-safercommunity-with-new-suicide-prevention-tools/ (accessed on 17 March 2019).

12. Dugas, A.F.; Hsieh, Y.H.; Levin, S.R.; Pines, J.M.; Mareiniss, D.P.; Mohareb, A.; Gaydos, C.A.; Perl, T.M.; Rothman, R.E. Google Flu Trends: Correlation with Emergency Department Influenza Rates and Crowding Metrics. Clin. Infect. Dis. 2012, 54, 463–469.

13. Ginsberg, J.; Mohebbi, M.H.; Patel, R.S.; Brammer, L.; Smolinski, M.S.; Brilliant, L. Detecting influenza epidemics using search engine query data. Nature 2009, 457, 1012–1014. [CrossRef]

14. Boland, K.M.; McNutt, J.G Assessing E-Government Success Strategies using Internet Search Data. In E-Government Success Factors Measures: Theories Concepts, Methodol; IGI Global: Hershey, PA, USA, 2013; pp. 1151–1169.

15. Bakker, K.M.; Martinez-Bakker, M.E.; Helm, B.; Stevenson, T.J. Digital epidemiology reveals global childhood disease seasonality and the e_ects of immunization. Proc. Natl. Acad. Sci. USA 2016, 113, 6689–6694.[CrossRef]

16. Pelat, C.; Turbelin, C.; Bar-Hen, A.; Flahault, A.; Valleron, A.-J. More Diseases Tracked by Using GoogleTrends. Clin. Infect. Dis. 2008, 47, 1443–1448.

17. Zhang, Y.; Milinovich, G.; Xu, Z.; Bambrick, H.; Mengersen, K.; Tong, S.; Hu, W. Monitoring PertussisInfections Using Internet Search Queries. Sci. Rep. 2017, 7, 10437

18. Rohart, F.; Milinovich, G.J.; Avril, S.M.R.; Lê Cao, K.; Tong, S.; Hu, W. Disease surveillance based on Internet-based linear models: An Australian case study of previously unmodeled infection diseases. Sci. Rep. 2016, 6, 38522.

19. Lampos, V.; Miller, A.C.; Crossan, S.; Stefansen, C. Advances in nowcasting influenza-like illness rates using search query logs. Sci. Rep. 2015, 5, 12760.

20. Cho, S.; Sohn, C.H.; Jo, M.W.; Shin, S.Y.; Lee, J.H.; Ryoo, S.M.; Kim, W.Y.; Seo, D.W. Correlation between national influenza surveillance data and google trends in South Korea. PLoS ONE 2013, 8, e81422.

21. Teng, Y.; Bi, D.; Xie, G.; Jin, Y.; Huang, Y.; Lin, B.; An, X.; Feng, D.; Tong, Y. Dynamic Forecasting of Zika Epidemics Using Google Trends. PLoS ONE 2017, 12, e0165085.

22. Dugas, A.F.; Jalalpour, M.; Gel, Y.; Levin, S.; Torcaso, F.; Igusa, T.; Rothman, R.E. Influenza forecasting with Google Flu Trends. PLoS ONE 2013, 8, e56176.

23. Towers, S.; Afzal, S.; Bernal, G.; Bliss, N.; Brown, S.; Espinoza, B.; Jackson, J.; Judson-Garcia, J.; Khan, M.; Lin, M.; et al. Mass Media and the Contagion of Fear: The Case of Ebola in America. PLoS ONE 2015, 10, e0129179.

24. Huang, D.C.; Wang, J.F. Monitoring hand, foot and mouth disease by combining search engine query data and meteorological factors. Sci. Total Environ. 2018, 612, 1293–1299.

25. Tenkanen, H.; di Minin, E.; Heikinheimo, V.; Hausmann, A.; Herbst, M.; Kajala, L.; Toivonen, T. Instagram, Flickr, or Twitter: Assessing the usability of social media data for visitor monitoring in protected areas. Sci. Rep. 2017, 7, 17615.

26. Reece, A.G.; Reagan, A.J.; Lix, K.L.M.; Dodds, P.S.; Danforth, C.M.; Langer, E.J. Forecasting the onset and course of mental illness with Twitter data. Sci. Rep. 2017, 7, 13006.

27. Shin, S.; Seo, D.; An, J.; Kwak, H.; Kim, S.; Gwack, J.; Jo, M. High correlation of Middle East respiratory syndrome spread with Google search and Twitter trends in Korea. Sci. Rep. 2016, 6, 32920

28. Thapen, N.; Simmie, D.; Hankin, C.; Gillard, J. DEFENDER: Detecting and Forecasting Epidemics Using Novel Data-Analytics for Enhanced Response. PLoS ONE 2016, 11, e0155417.

29. Allen, C.; Tsou, M.; Aslam, A.; Nagel, A.; Gawron, J. Applying GIS and Machine Learning Methods toTwitter Data for Multiscale Surveillance of Influenza. PLoS ONE 2016, 11, e0157734.

30. Volkova, S.; Ayton, E.; Porterfield, K.; Corley, C.D. Forecasting influenza-like illness dynamics for military populations using neural networks and social media. PLoS ONE 2017, 12, e0188941

31. Simon, T.; Goldberg, A.; Aharonson-Daniel, L.; Leykin, D.; Adini, B. Twitter in the Cross Fire—The Use of Social Media in the Westgate Mall Terror Attack in Kenya. PLoS ONE 2014, 9, e104136. [CrossRef] [PubMed]

32. Xia, F.; Su, X.; Wang, W.; Zhang, C.; Ning, Z.; Lee, I. Bibliographic Analysis of Nature Based on Twitter and Facebook Altmetrics Data. PLoS ONE 2016, 11, e0165997.

33. Patel, R.; Belousov, M.; Jani, M.; Dasgupta, N.; Winakor, C.; Nenadic, G.; Dixon, W.G. Frequent discussion of insomnia and weight gain with glucocorticoid therapy: An analysis of Twitter posts. Npj Digit. Med. 2018, 1, 7

34. Xu, Q.; Gel, Y.R.; Ramirez Ramirez, L.L.; Nezafati, K.; Zhang, Q.; Tsui, K.L. Forecasting influenza in Hong Kong with Google search Queries and statistical model fusion. PLoS ONE 2017, 12, e0176690

35. He, F.; Hu, Z.; Zhang, W.; Cai, L.; Cai, G.; Aoyagi, K. Construction and evaluation of two computational models for predicting the incidence of influenza in Nagasaki Prefecture, Japan. Sci. Rep. 2017, 7, 7192.

36. Najafabadi, M.M.; Villanustre, F.; Khoshgoftaar, T.M.; Seliya, N.; Wald, R.; Muharemagic, E. Deep learning applications and challenges in big data analytics. J. Big Data 2017, 2.

37. Janowczyk, A.; Madabhushi, A. Deep learning for digital pathology image analysis: A comprehensive tutorial with selected use cases. J. Pathol. Inform. 2016, 7, 29.

38. Esteva, A.; Kuprel, B.; Novoa, R.A.; Ko, J.; Swetter, S.M.; Blau, H.M.; Thrun, S. Dermatologist-level classification of skin cancer with deep neural networks. Nature 2017, 542, 115.

39. Bychkov, D.; Linder, N.; Turkki, R.; Nordling, S.; Kovanen, P.E.; Verrill, C.; Walliander, M.; Lundin, M.; Haglund, C.; Lundin, J. Deep learning based tissue analysis predicts outcome in colorectal cancer. Sci. Rep. 2018, 8, 3395.

40. Aldhyani THH, Alshebami AS, Alzahrani MY. Soft Computing Model to Predict Chronic Diseases. Journal of Information Science and Engineering. 36(2). 2020, 365–376 10.6688/JISE.202003

41. Aldhyani THH, Alshebami AS, Alzahrani MY. Soft Clustering for Enhancing the Diagnosis of Chronic Diseases over Machine Learning Algorithms. J Healthc Eng. 2020;. Published 2020 Mar 9. 2020:4984967 doi:10.1155/2020/4984967

42. D. Balcan, V. Colizza, B. Goncalves, H. Hu, J. Ramasco and A. Vespignani, “Multiscale mobility networks and the spatial spreading of infectious diseases” Proceedings of the National Academy of Sciences, vol. 106, no. 51, pp. 21484–21489, 2009. Available: 10.1073/pnas.0906910106.

43. Q. Yuan, E. Nsoesie, B. Lv, G. Peng, R. Chunara and J. Brownstein, “Monitoring Influenza Epidemics in China with Search Query from Baidu” PLoS ONE, vol. 8, no. 5, p. e64323, 2013. Available: 10.1371/journal.pone.0064323.

44. Milinovich GJ, Avril SM, Clements AC, Brownstein JS, Tong S, Hu W. Using internet search queries for infectious disease surveillance: screening diseases for suitability. BMC Infect Dis 2014;14:690 [FREE Full text] [doi: 10.1186/s12879-014-0690-1] [Medline: 25551277]

45. G. Milinovich, S. Avril, A. Clements, J. Brownstein, S. Tong and W. Hu, “Using internet search queries for infectious disease surveillance: screening diseases for suitability” BMC Infectious Diseases, vol. 14, no. 1, 2014. Available: 10.1186/s12879-014-0690-1

46. S. Cook, C. Conrad, A. Fowlkes and M. Mohebbi, “Assessing Google Flu Trends Performance in the United States during the 2009 Influenza Virus A (H1N1) Pandemic” PLoS ONE, vol. 6, no. 8, p. e23610, 2011. Available: 10.1371/journal.pone.0023610.

47. https://www.kaggle.com/sudalairajkumar/novel-corona-virus-2019-dataset.

48. Qiang Zhang, Tianse Gao, Xueyan Liuand Yun Zheng. Public “Environment Emotion Prediction Model Using LSTM Network”. Sustainability 2020, 12, 1665;2020

